# Duchenne muscular dystrophy patients lacking the dystrophin isoforms Dp140 and Dp71 and mouse models lacking Dp140 have a more severe motor phenotype

**DOI:** 10.1101/2021.07.27.21261120

**Authors:** Mary Chesshyre, Deborah Ridout, Yasumasa Hashimoto, Yoko Ookubo, Silvia Torelli, Kate Maresh, Valeria Ricotti, Lianne Abbott, Vandana Ayyar Gupta, Marion Main, Mariacristina Scoto, Giovanni Baranello, Adnan Manzur, Yoshitsugu Aoki, Francesco Muntoni, On behalf of the North Star clinical network

## Abstract

**Background:** Duchenne muscular dystrophy (DMD) is caused by *DMD* mutations leading to dystrophin loss. Full length Dp427 is the primary dystrophin isoform expressed in skeletal muscle and is also expressed in the central nervous system (CNS). Two shorter isoforms, Dp140 and Dp71, are highly expressed in the CNS. While a role for Dp140 and Dp71 on DMD CNS co-morbidities is well known, relationships between lack of Dp140 and Dp71 and DMD motor outcomes are not. We have conducted a series of investigations addressing this.

**Methods:** Functional outcome data from 387 DMD boys aged 4.0-15.4 years was subdivided by *DMD* mutation expected effect on isoform expression; Group 1 (Dp427 absent, Dp140/Dp71 present, n=201); group 2 (Dp427/Dp140 absent, Dp71 present, n=152); and group 3 (Dp427/Dp140/Dp71 absent, n=34). Relationships between isoform group and North Star ambulatory assessment (NSAA) scores, 10m walk/run and rise times were explored using regression analysis. We used Capillary Western immunoassay (Wes) analysis to study Dp427, Dp140 and Dp71 production in wild-type and DMD skeletal muscle and myogenic cultures. Grip strength was studied in wild-type, *mdx* (Dp427 absent, Dp140/Dp71 present), *mdx52* (Dp427/Dp140 absent, Dp71 present) and *DMD-null* (lacking all isoforms) mice.

**Results:** In DMD boys, we found a strong association between isoform group and motor function. In DMD boys, mean NSAA scores at 5 years of age were 6.1 points lower in group 3 than group 1 (p<0.01) and 4.9 points lower in group 3 than group 2 (p=0.05). Mean peak NSAA scores were 4.0 points lower in group 3 than group 1 (*p*<0.01), 2.4 points lower in group 3 than group 2 (*p*=0.09) and 1.6 points lower in group 2 than group 1 (*p*=0.04).

Average grip strength in peak force at 3 months of age was higher in *mdx* than *mdx52* mice (*p*=0.01).

Dp427, but not Dp71, was produced in normal skeletal muscle; low levels of Dp71 were detected in DMD skeletal muscle. High Dp71 levels were present in wild-type and DMD myogenic cultures.

**Conclusions:** DMD boys lacking Dp140 and Dp140/Dp71 displayed worse motor function with a cumulative effect of isoform loss. DMD mouse models lacking Dp427 and Dp140 had lower grip strength than those lacking Dp427 but not Dp140. Our results highlight the importance of considering the effects of dystrophin isoform loss on DMD motor impairment, with important implications for understanding the complex relationship between brain and muscle function in DMD and patient stratification for clinical trials.

## Introduction

Duchenne muscular dystrophy (DMD) is an X-linked recessive condition caused by *DMD* mutations leading to deficiency of dystrophin[1,2]

Boys with DMD typically present with delayed motor milestones, frequent falls and speech delay[2]. Neurobehavioural comorbidities, including intellectual disability, attention deficit hyperactivity disorder and/or autism spectrum disorder, occur in approximately one third of patients[2]. Loss of ambulation typically occurs by 12 years of age, followed by progression to cardiomyopathy and respiratory insufficiency[2]. Mean age of survival is in the late twenties[2].

DMD shows significant clinical heterogeneity. Some heterogeneity is related to *DMD* genotypes allowing production of low levels of dystrophin[3]. In addition, changes in genes other than *DMD*, that modify the clinical course, have recently been identified[4–6]. However, each of these modifiers contributes only modestly to DMD trajectory, suggesting the existence of other factors.

The *DMD* locus encodes for multiple isoforms[7]. Full-length Dp427 exists in 3 almost identical isoforms. Dp427m, is the isoform most highly expressed in skeletal muscle. Dp427c and Dp427p are expressed in the cortex, amygdala, hippocampus and cerebellum[7]. Additional shorter dystrophin isoforms are driven by promoters in introns further downstream the gene. Two of which, Dp140 and Dp71, are abundantly expressed in the brain[7,8]. Whilst in DMD, all mutations affect Dp427 production, the *DMD* mutation can also result in disruption of one or both of Dp140 and Dp71. *DMD* mutations involving the genomic region upstream of intron 44 only affect Dp427; mutations involving the genomic region from exon 51 to exon 62 inclusive, but not involving the genomic region of exon 63 and/or the genomic region downstream of exon 63, also affect Dp140, but not Dp71, and mutations involving the genomic region of exon 63 and/or the genomic region downstream of exon 63 affect Dp427, Dp140 and Dp71[7,8]. The effect of mutations involving the genomic region from exon 45 to exon 50 inclusive and not involving the genomic region of exon 51 and/or the genomic region downstream of exon 51 on Dp140 production cannot be predicted precisely, as the Dp140 promoter is located in intron 44 and its translation start site is located in exon 51[8,9]. Other isoforms also exist (Dp260 and Dp116), with patterns of expression limited to the retina and peripheral nerves respectively[7].

Several studies have suggested a role of Dp140 and Dp71 in DMD central nervous system (CNS) involvement[10–13]. In a European DMD cohort, 15% of boys lacking only Dp427 had intellectual disability, compared with 25% of boys lacking Dp427 and Dp140 and 64% of boys lacking Dp427, Dp140 and Dp71[13].

We hypothesised that *DMD* mutations affecting different dystrophin isoforms may have a different impact on DMD motor outcomes. We conducted a series of studies to address this. We evaluated the relationship between mutations leading to loss of Dp427/Dp140 and Dp427/Dp140/Dp71 compared to loss of Dp427 alone on motor function in a large cohort of DMD boys. We focused on four motor outcomes commonly used in clinical trials: (1) North Star ambulatory assessment (NSAA) score, (2) rise from supine time and velocity, (3) 10 metre walk/run time and velocity, and (4) age at loss of ambulation (LOA) in boys with DMD. We evaluated grip strength in wild-type and 3 DMD mouse models with mutations associated with different patterns of dystrophin isoform expression. Finally, we evaluated dystrophin isoform protein production in wild type and DMD skeletal muscle and myogenic cells in culture.

## Methods

### Standard protocol, approvals, registrations, and patient consents

The Dubowitz Neuromuscular Centre coordinates the North Star network of 23 UK neuromuscular centres looking after DMD patients[13–15]. Clinical data is prospectively collected in routine clinical appointments every six months. There is a national training programme and standard operating procedures for each site. Clinical data is stored in an electronic database managed by CertusLtd[13]. Participants meeting inclusion criteria were recruited from this network.

### Study design and participants

387 participants aged 4.0-15.4 years, with a genetically confirmed DMD diagnosis or a diagnosis of intermediate muscular dystrophy (IMD) and an out-of-frame *DMD* deletion or duplication were included. Participants with in-frame *DMD* deletions/duplications and an IMD phenotype, BMD, manifesting carriers, clinical trial participants and boys under 4 years were excluded.

### NorthStar natural history study data

Clinical data reviewed from the NorthStar natural history study included *DMD* mutation, date of DMD diagnosis, NSAA scores, rise from supine time, 10 metre walk/run time, age of LOA and presence or absence of learning disabilities. The NSAA is a 17-item DMD-specific motor function scale with a maximum possible total score of 34[16].

For the functional data, we analysed data at 5 years of age, as this is the standard lower end of age at recruitment in most clinical trials; the age of peak functional achievement, and the age at loss of ambulation.

### *DMD* mutation data

Participants were grouped into 3 groups based on predicted *DMD* mutation effects on dystrophin isoform expression: Group 1 (Dp427 absent, Dp140/Dp71 present, n=201); group 2 (Dp427/Dp140 absent, Dp71 present, n=152); and group 3 (Dp427/Dp140/Dp71 absent, n=34). Based on the genomic organisation of the *DMD* locus[7,8], patients with mutations only involving the genomic region upstream of intron 44 were considered to be Dp427 negative and Dp140/Dp71 positive (group 1), patients with mutations involving the genomic region from exon 51 to exon 62 inclusive, but not involving the genomic region of exon 63 and/or the genomic region downstream of exon 63 were considered Dp427/Dp140 negative and Dp71 positive (group 2) and patients with mutations involving the genomic region of exon 63 and/or the genomic region downstream of exon 63 were considered to be Dp427/Dp140/Dp71 negative (group 3)[8,9]. Patients with mutations involving the genomic region from exon 45 to exon 50 inclusive and not involving the genomic region of exon 51 and/or the genomic region downstream of exon 51 were excluded from analysis due to the difficulty in predicting the effects of these mutations on Dp140 expression, as the Dp140 promoter is located in intron 44 and its translation start site is located in exon 51[8,9].

### Cognitive status

Cognitive status was evaluated in a subset of boys by both formal testing of intelligence quotient (IQ) and presence or absence of learning difficulties. Presence or absence of learning difficulties was collected for the NorthStar natural history study and determined by parental and/or educational report.

Formal IQ testing was carried out in a subset of 40 boys who have participated in other studies undertaken at our centre - Ricotti et al[10] and Maresh et al (manuscript in preparation). Methods of IQ assessments are reported in our previous publication[10].

Participants were categorised into two groups based on cognitive status. The cognitive impairment group (n=56) had learning disabilities (as reported by parent/guardian(s) and/or educational services) and/or an IQ of less than 85 (>1 standard deviation below the mean population IQ)[17,18]. The normal cognition group (n=71) had no learning disability (as reported by parent/guardian(s) and/or educational services) and/or an IQ of 85 or above.

### Statistical analysis for analysis in DMD boys

Patient characteristics were summarised using mean and standard deviation for continuous data and frequency and proportion for categorical data. GC regimen was summarised as the regime (Daily, Intermittent/Other or None) taken for the longest duration over both the full longitudinal period (majority regimen) and the duration prior to peak NSAA (early regime). Where age of GC initiation was not available (139/340 of boys on GC), this was estimated based on mean age of GC initiation in the cohort. Ambulatory function at 5 years was described for patients for whom a visit was recoded between 4.5-5.5 years of age.

Comparisons between isoform groups and neurocognitive impairment groups were made using one-way analysis of variance and Chi-squared tests. For the 3 main longitudinal outcomes, NSAA total score, 10m walk/run velocity and rise time velocity, relationships with age were explored for each isoform group; additionally, the relationship for NSAA total score with age was explored in the 2 cognition groups. Fractional polynomial regression, accounting for the longitudinal data, was used to find best fitting models, by comparing model deviances^22^. Noting the different observed mean peak functional scores for the dystrophin isoform groups and neurocognitive groups, we focused subsequent analysis on the age range 5.5–8 years, where we expected the majority of participants to reach peak motor function. Within this age interval, we calculated maximum observed motor function scores and age of maximum attainment for the 3 main outcomes.

Multivariable regression analysis was used to compare maximum scores achieved between dystrophin isoform groups and neurocognitive groups. We assessed whether differences in the majority GC regimen used up to the time of maximum score accounted for differences between dystrophin isoform groups. Kaplan-Meier survival estimation was used to estimate median time to LOA for each dystrophin isoform group. *P* value less than or equal to 0.05 was considered statistically significant.

Statistical analysis and creation of figures was conducted by Dr Deborah Ridout in Stata v 15 StataCorp. 2017. *Stata Statistical Software: Release 15*. College Station, TX: StataCorp LLC

### Mouse DMD models

All mice used in this study were maintained at the National Center of Neurology and Psychiatry (NCNP)[19]. 3 different dystrophic mouse models were studied, with differential dystrophin isoform involvement; *mdx* mice (Dp427 absent, Dp140/Dp71 present), *mdx-52* mice (Dp427/Dp140 absent, Dp71 present) and *DMD-null* mice lacking all dystrophin isoforms[20–22]. *Mdx52* mice were kindly provided by Dr T. Sasaoka (Brain Research Institute, Niigata University, Niigata, Japan)[23]. *DMD-null* mice were generated by Dr Kazunori Hanaoka[22]. C57BL/6 mice were used as controls to match the background of *mdx, mdx52* and *DMD-null* mice. Genotyping was performed using previously described PCR method[21–23]. Animal care was provided by the Small Animal Research Facility at the NCNP. Mice were allowed ad libitum access to food and drinking water. All behavioural experiments were performed between 9 am and 1 pm in strict accordance with the regulations of the National Institute of Neuroscience and the National Center of Neurology and Psychiatry (Japan) for animal experiments and were approved by the Animal Investigation Committee of the Institute. Statistical analysis and figures were carried out using GraphPad Prism8 (GraphPad Software Inc., La Jolla, CA).

### Mouse forelimb grip strength test

The grip strength test was conducted on male dystrophic mouse models, including *mdx* (n=9), *mdx52* (n=10), *DMD-null* (n=3) mice at 3 months of age. Mice were tested to determine peak paw grip strength, positioned horizontally from a grip bar using a grip strength meter (MK-380 M; Muromachi Kikai Co., Ltd., Japan) as previously described, and pulled back slowly and steadily until these mice released their grip[24]. This was repeated 6 times, and peak force for the forelimb paws was measured. These mice weighed on the date of their grip strength testing to be normalized by their body weight. The degree of grip fatigue (Force-decline rate) is calculated by comparing the first two pull to the last two pull. In the formula (5^th^ +6^th^) / (1^st^ +2^nd^) *100 (%) gives a measure of fatigue.

### Dystrophin isoform protein production in wild type and DMD skeletal muscle and myogenic cells

All patients and controls provided written informed consent for skin and muscle biopsy samples. Fibroblasts and muscle tissues were supplied by the MRC Centre for Neuromuscular Disease Biobank London.

Tissue sampling was approved by the NHS national Research Ethics Service as follows under the studies ‘Setting up of a rare diseases biological samples bank (biobank) for research to facilitate pharmacological, gene and cell therapy trials in neuromuscular disorders (NMD)’ (REC reference number: 06/Q0406/33 - Hammersmith and Queen Charlotte’s and Chelsea Research Ethics Committee, ‘The use of cells as a model system to study pathogenesis and therapeutic strategies for Neuromuscular Disorders’ (REC reference 13/LO/1826 - London - Stanmore Research Ethics Committee) and ‘Genes and Proteins in Neuromuscular Disorders’ (REC reference 13/LO/1894 - London - Camberwell St Giles Research Ethics Committee).

Proteins were extracted from human and DMD dermal fibroblasts differentiated into myotubes after transfection by a lentiviral-mediated myoD construct carrying a puromycin selection cassette (for transduced cell enrichment) and a DsRed cassette (for assessing transduction efficiency). Protein lysates from control and DMD muscle were prepared from snap frozen tissues. Protein concentrations were measured using the Pierce BCA Protein assay kit (Thermo Scientific 23225), according to the manufacturer’s instructions.

Capillary Western immunoassay (Wes) analysis was performed on a Wes system (ProteinSimple) according to the manufacturer’s instructions using a 66–440 kDa Separation Module (ProteinSimple). For dystrophin a rabbit polyclonal anti-dystrophin antibody (ab15277, Abcam, dilution 1/50) and an anti-rabbit secondary antibody (042-206, Protein Simple) were used. The following amount of protein lysate (μg per well/capillary) were loaded for each sample: 1µg control fibroblasts MyoD-transfected; 1µg DMD fibroblasts MyoD-transfected; 0.125µg control muscle; 0.250μg DMD muscle.

## Results

### Patient characteristics

Participants lacking both the Dp427 and Dp140 isoforms, but not Dp71 (group 2) were on average diagnosed with DMD 6 months earlier than those who only lacked Dp427 (group 1, *p*=0.05, Table 1). There was no difference between groups 1 and 3 (*p*=0.99) and 2 and 3 (*p*=0.99).

**Table 1.** **Patient characteristics: DMD=Duchenne muscular dystrophy, sd=standard deviation, NSAA=North Star ambulatory assessment, G1=group 1, G2=group 2, G3=group 3, GC=glucocorticoid, na = not estimable.**

|  | Group 1<br>(n=201)<br>Mean (sd) | Group 2<br>(n=152)<br>Mean (sd) | Group 3<br>(n=34)<br>Mean (sd) | All patients<br>(n=387)<br>Mean (sd) | P value<br>between<br>groups |
| --- | --- | --- | --- | --- | --- |
| **Demographics** |  |  |  |  |  |
| Age at diagnosis of DMD<br>(years)<br>G1 n=171, G2 n=123, G3<br>n=31 | 4.2 (2.0) | 3.6 (2.0) | 3.8 (2.0) | 3.9 (2.0) | 0.05 |
| Age range at baseline<br>(years) | 4.0, 15.4 | 4.0, 12.0 | 4.0, 12.2 | 4.0, 15.4 |  |
| **GC Use** |  |  |  |  |  |
| Age at initiation (years)<br>G1=94, G2=71, G3=12 | 5.8 (1.4) | 5.5 (1.3) | 6.0 (1.5) | 5.7 (1.5) | 0.22 |
| GC use recorded at any<br>time (n and %) | 189 (94.0<br>%) | 147<br>(96.7%) | 31<br>(91.2%) | 367 (94.8%) | 0.32 |
| Majority Regime<br>Daily (n and %)<br>Int/Other (n and %)<br>None (n and %) | 94 (46.8%)<br>85 (42.3%)<br>22 (10.9%) | 78 (51.3%)<br>58 (38.2%)<br>16 (10.5%) | 13 (38.2%)<br>12 (35.3%)<br>9 (26.5%) | 185 (47.8%)<br>155 (40.1%)<br>47 (12.1%) | 0.09 |
| **Loss of Ambulation** |  |  |  |  |  |

|  | **Group 1**<br>**(n=148)** | **Group 2**<br>**(n=108)** | **Group 3**<br>**(n=23)** | **All patients**<br>**(n=279)\*** | **P value**<br>**between**<br>**groups** |
| --- | --- | --- | --- | --- | --- |
| Median (IQR) age of loss of ambulation (years) | 15.7<br>(11.7, na) | 13.0<br>(12.1, 14.9) | 13.6<br>(12.9, 14.1) | 13.6 (11.8, 16.1) | p=0.67 |
| **Ambulatory Function at 5 years of age** |  |  |  |  |  |
| | **Group 1**<br>**(n=44)**<br>**Mean (sd)** | **Group 2**<br>**(n=37)**<br>**Mean (sd)** | **Group 3**<br>**(n=9)**<br>**Mean (sd)** | **All patients**<br>**(n=90)\$**<br>**Mean (sd)** | **P value**<br>**between**<br>**groups** |
| NSAA score at 5 years of age | 22.6 (5.4) | 21.4 (5.0) | 16.5 (6.7) | 21.5 (5.6) | 0.01 |
| Rise from supine time at 5 years of age (seconds)<br>G1 n=40, G2 n=37, G3 n=9 | 4.9 (2.1) | 4.9 (2.0) | 9.5 (11.5) | 5.4 (4.3) | <0.01 |
| 10m walk/run time at 5 years of age (seconds)<br>G1 n=41, G2 n=29, G3 n=9 | 6.7 (2.6) | 6.6 (2.4) | 7.0 (2.2) | 6.7 (2.5) | 0.90 |

\$90 patients had a NSAA recorded between ages 4.5-5.5 years

We did not find a statistically significant difference in age of GC initiation (p=0.22), GC regime (*p*=0.09) or GC use recorded at any time between dystrophin isoform group, table 1. There was a non-significant trend for those in group 3 to be more likely to be on no GC (p=0.09, table 1).

### Ambulatory function in DMD boys at 5 years of age

Mean NSAA score at 5 years of age was over 6 points lower in participants lacking all isoforms (Dp427/Dp140/Dp71; group 3) compared to those only lacking Dp427 (group 1; *p*<0.01) and 4.9 points lower in group 3 than those lacking both Dp427 and Dp140, but able to express Dp71 (group 2) (*p*=0.05, Table 1). There was no difference between groups 1 and 2 (*p*=0.99).

Mean rise from supine times at 5 years of age were 4.6 seconds slower in group 3 than group 1 (*p*<0.01) and 4.6 seconds slower in group 3 than group 2 (*p*=0.01, Table 1). There was no difference between groups 1 and 2 (*p*=0.99).

There was no difference in mean 10m walk/run time at 5 years of age between dystrophin isoform groups (p=0.99 for all group-wise comparisons). 10m walk/run time can be considered to be a less complex assessment than rise from supine time and the NSAA.

These results suggest that at 5 years of age, lack of Dp71 in addition to Dp427 and Dp140 was associated with poorer motor function in the two more complex motor assessments.

### Peak motor function in DMD boys in the dystrophin isoform groups

Previous studies have reported a mean peak in NSAA score at 6-7 years of age[14,15]. In our NSAA score trajectory models, peak NSAA scores in the different isoform groups occurred within the age range of 5.5-8.0 years (Fig 1). Therefore, to assess if the isoform groups differed in maximal functional achievements, we looked at differences in peak mean NSAA score, 10m walk/run velocity and rise from supine time in the observed data for those aged 5.5-8.0 years in the dystrophin isoform groups (Table 2).

**Table 2.** **Mean peak NSAA scores for the dystrophin isoform groups overall and stratified by GC regime, mean peak 10m walk/run velocity and mean peak rise from supine time velocities not stratified by GC regimen. G1 = group 1, G2 = group 2, G3 = group 3 and GC=glucocorticoid. This table considers the dataset for a subset (n=263) of boys, aged 5.5 – 8.0 years, for whom majority GC regime data and peak NSAA scores were available for all 262 boys. This subset of 5.5-8.0 years of age was used as this is the age in which the majority of participants reach peak motor function[14,15]**

|  | Group 1<br>n=129 | Group 2<br>n=114 | Group 3<br>n=20 | All groups<br>n=263 |
| --- | --- | --- | --- | --- |
| **Early Regime (prior to peak NSAA)** | 56 (43.4) | 41 (36.0) | 7 (35.0) | 104 (39.5) |
| **Daily** | 55 (42.6) | 59 (51.7) | 7 (35.0) | 121 (46.0) |
| **Int/Other** | 18 (14.0) | 14 (12.3) | 6 (30.0) | 38 (14.5) |
| **None** |  |  |  |  |
| **Mean (sd) peak NSAA score for those on daily GC** | 26.9 (5.1) | 26.4 (4.3) | 24.1 (4.8) | 26.5 (4.8) |
| **Mean (sd) peak NSAA score for those on intermittent or other GC** | 26.8 (5.7) | 24.6 (6.6) | 24.1 (5.0) | 25.6 (6.2) |
| **Mean (sd) peak NSAA score for those on no GC** | 24.9 (6.4) | 22.3 (6.3) | 17.5 (10.0) | 22.8 (7.3) |
| **Mean (sd) peak NSAA score overall (all GC regimens combined)** | 26.6 (5.5) | 25.0 (5.9) | 22.1 (7.1) | 25.6 (5.9) |
| **Mean (sd) peak 10m walk/run velocity overall (m/s)** | 2.1 (0.5)<br>n=121 | 1.9 (0.6)<br>n=108 | 1.7 (0.5)<br>n=18 | 2.0 (0.6)<br>n=247 |
| **Mean (sd) peak rise from supine time velocity overall (m/s)** | 0.29 (0.12)<br>n=126 | 0.28 (0.13)<br>n=105 | 0.25 (0.12)<br>n=17 | 0.28 (0.12)<br>n=248 |

**Figure 1.**
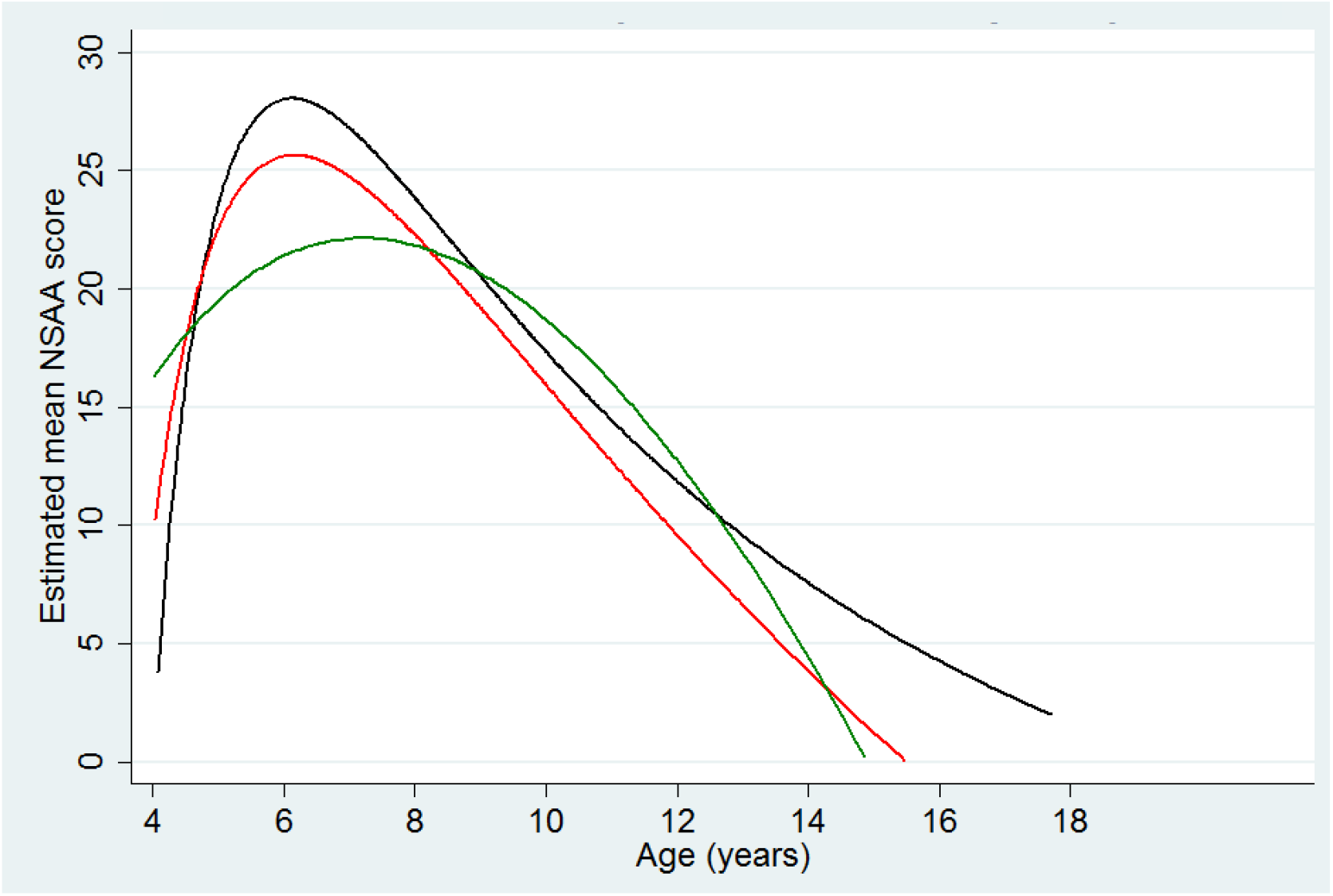
**Estimated mean NSAA score trajectory with age models in the dystrophin isoform groups. Each line represents estimated mean NSAA score plotted against age for the dystrophin isoform group. Group 1 = blue, group 2 = red, group 3 = green.**

Mean peak NSAA scores were progressively lower for groups lacking more dystrophin isoforms; 4.0 points lower in group 3 than group 1 (*p*<0.01), 2.4 points lower in group 3 than group 2 (*p*=0.09) and 1.6 points lower in group 2 than group 1 (*p*=0.04; Table 2), after adjusting for GC regime. The age at attainment of peak score was not significantly different between the isoform groups (*p*=0.10).

Similarly, mean peak 10m walk/run velocity was 0.4 m/s (95%CI 0.2-0.7) slower in group 3 than group 1 (*p*<0.001) and 0.2 m/s (95%CI (0.05-0.4) slower in group 2 than group 1 (*p*<0.001), and non-significantly lower in group 3 compared to group 2 (*p*=0.10), after adjusting for GC regime. There was no difference in age of mean peak 10m walk/run velocity between isoform groups (p=0.91). Peak rise from supine time velocity was 0.04 m/s slower in group 3 than group 1 and 0.03 m/s slower in group 3 than group 2, however these differences were not significant (p=0.61).

### Age of loss of ambulation (LOA) in DMD boys

The median ages of loss of ambulation were lower in groups 2 and 3 than group 1 (15.7 years in group 1, 13.0 years in group 2 and 13.6 years in group 3), however these differences were not significant (Table 1, p=0.67).

### Effects of dystrophin isoform group on functional trajectories

#### NorthStar ambulatory assessment trajectory stratified by dystrophin isoform group

In NSAA score trajectory models, mean NSAA scores were lower in group 2 than group 1 (Fig 1). From 4.5-8.5 years of age, mean NSAA scores were lower in group 3 than in groups 1 and 2 with a cumulative effect of loss of isoforms (Fig 1). Peak estimated NSAA scores were lower in groups 2 and 3 with a cumulative effect of loss of isoforms (Fig 1).

### 10m walk/run velocity trajectory model stratified by dystrophin isoform group

In models of 10m walk/run velocity trajectory with age, velocities were higher in group 1 than groups 2 and 3 with a cumulative effect of loss of isoforms from approximately 6-8 years of age (Fig 2).

**Figure 2.**
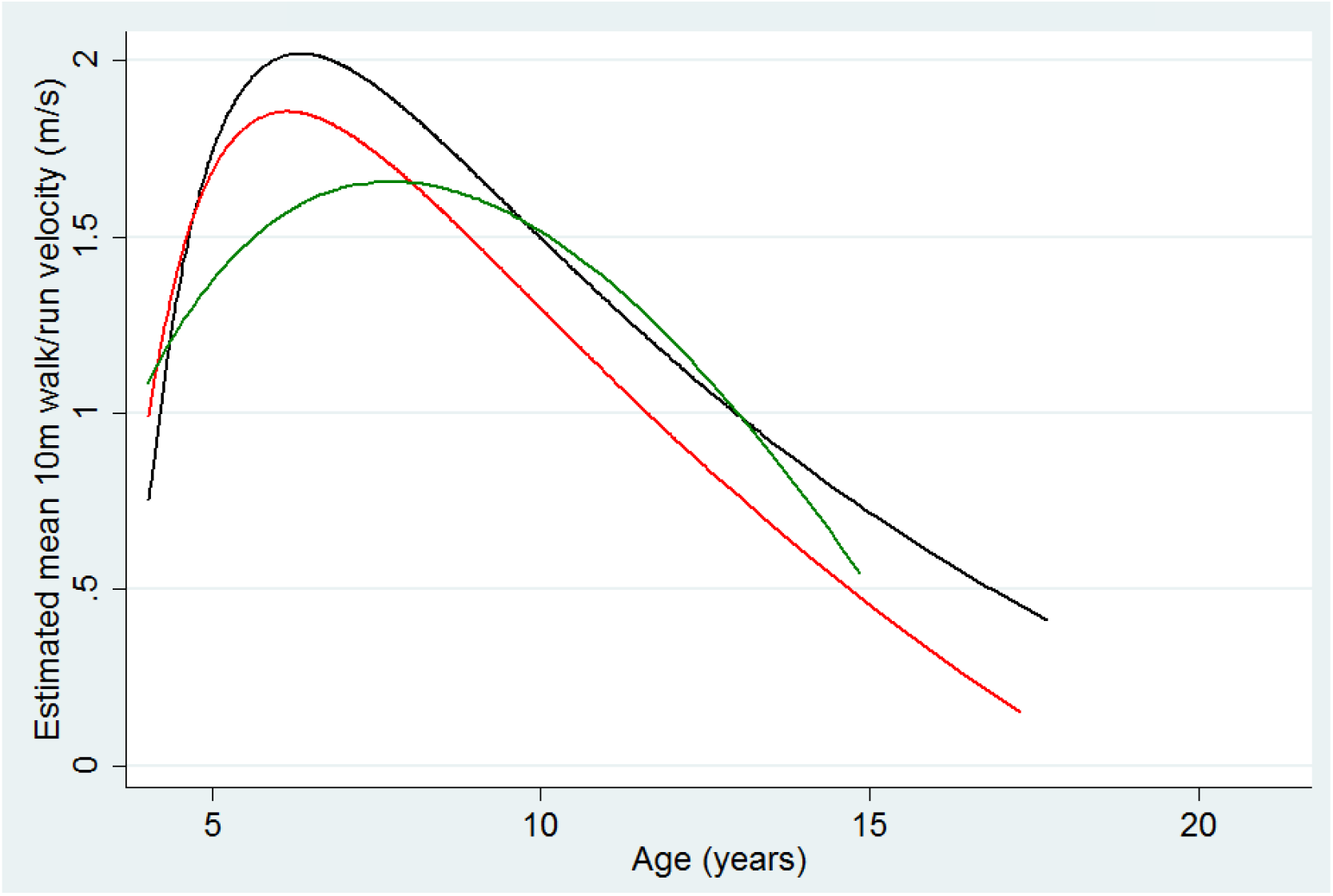
**Estimated mean 10m walk/run velocity trajectory models in the dystrophin isoform groups. Each line represents estimated mean NSAA score trajectory plotted against age for the dystrophin isoform group. Group 1 = blue, group 2 = red, group 3 = green.**

### Rise from supine time velocity trajectories stratified by dystrophin isoform group

In mean rise from supine time velocity (m/s) trajectory models, group 3 had a slower estimated mean rise from supine time velocity than groups 1 and 2 from approximately 5-9 years of age (Fig 3).

**Figure 3.**
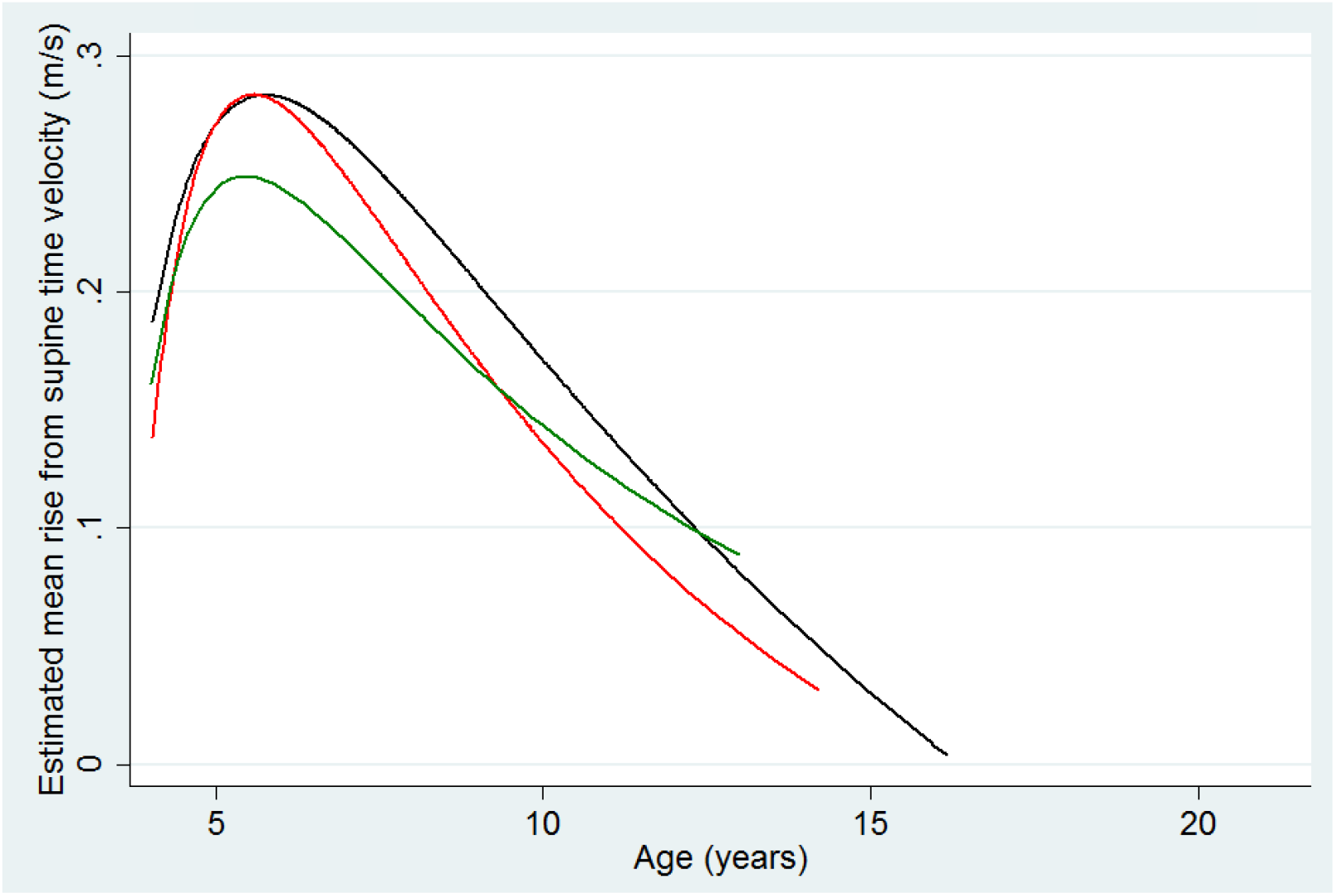
**Estimated mean rise from supine time velocity trajectory models in the dystrophin isoform groups. Each line represents estimated mean NSAA score trajectory plotted against age for the dystrophin isoform group. Group 1 = blue, group 2 = red, group 3 = green.**

### Influence of cognitive impairment on functional outcomes

We evaluated the influence of cognitive impairment on peak NSAA scores in a subset of 127 boys for whom cognition group, GC regimen and peak NSAA scores were available.

We first confirmed a strong cumulative association between lack of brain dystrophin isoforms and cognitive impairment (p<0.001, Supplementary Table 1). Cognitive impairment was found in 32% (20/63) of those in group 1, 53% (29/55) of those in group 2 and 78% (7/9) of those in group 3 (Supplementary Table 1), in keeping with previously published studies[10–13].

Mean peak NSAA scores were 2.2 points lower in those with cognitive impairment than those with normal cognition (*p*=0.04), after adjusting for GC regimen (Supplementary table 1).

In NSAA trajectory models stratified by cognitive group, mean NSAA scores were lower in those with cognitive impairment than those without cognitive impairment at all ages (Fig 4)

**Figure 4.**
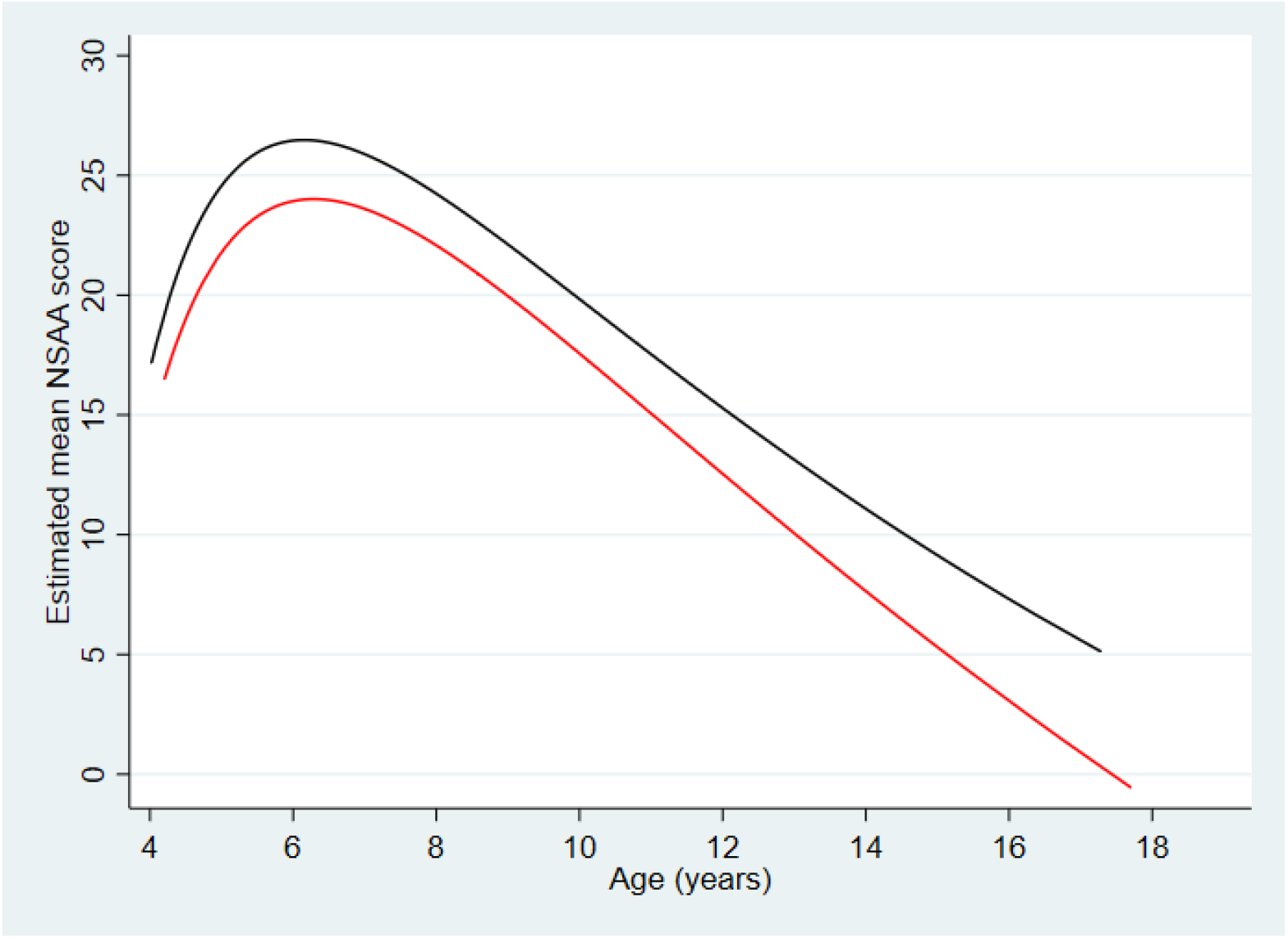
**Estimated mean NSAA score trajectory with age models in those with and without cognitive impairment. Each line represents estimated mean NSAA score trajectory plotted against age for the cognition group. Black = normal cognition and red = impaired cognition.**

### Forelimb grip strength measurements in *mdx, mdx52* and *DMD-null* mice at the age of 3 months

Average grip strength in peak force at 3 months of age was significantly higher for *mdx* mice than that of *mdx52* mice (*p* = 0.01), with statistical differences between WT and *mdx52* mice, and between WT and *DMD-null* mice (Fig. 5a). There were no statistically significant differences in the degree of grip fatigue at the age of 3 months between the three DMD mouse strains.

**Figure 5.**
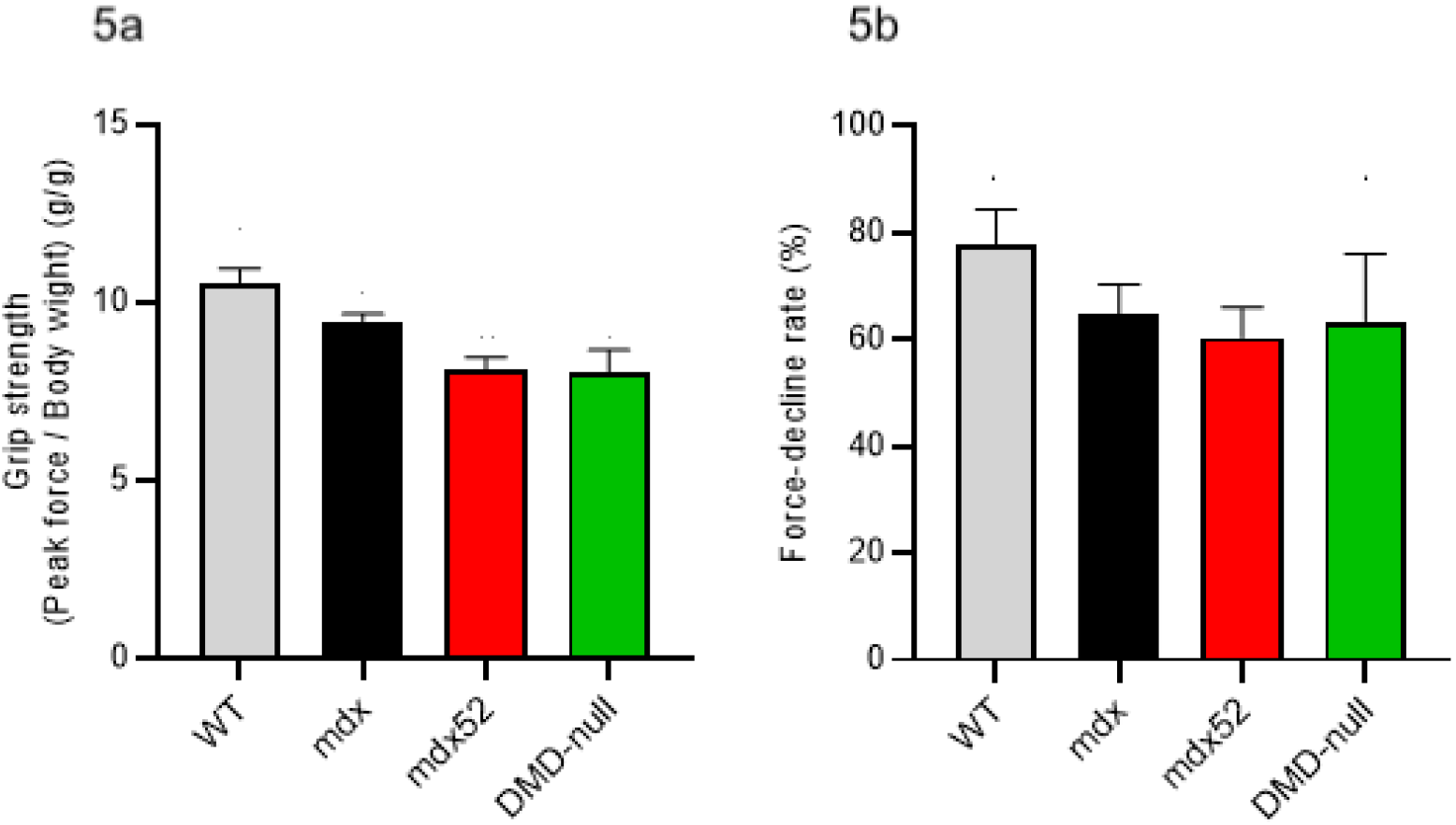
**Repeat testing of grip strength in WT (n=4), *mdx* (n=9), *mdx52* (n=10) and *DMD-null* (n=3) mice at the age of 3 months. Mean grip strength at the age of 3 months; *n =* 3-10, Mean ± SEM, One-way ANOVA with Dunnett *p* = 0.0005 (WT vs *mdx52*), *p* = 0.0052 (WT vs *DMD-null*), *p =* 0.01 (*mdx* vs *mdx52*). (5a). The degree of grip fatigue at the age of 3 months. Mean ± SEM, One-way ANOVA with Dunnett (5b). There was no statistically significant difference in grip fatigue at the age of 3 months between the 3 DMD mouse strains.**

### Dystrophin isoform protein production in wild type and DMD skeletal muscle and myogenic cells

A band corresponding to Dp71 was seen in control and DMD MyoD transfected fibroblasts and in DMD skeletal muscle, but not in control skeletal muscle (Fig 6). The band corresponding to full length dystrophin was only seen in MyoD transfected control fibroblasts and control skeletal muscle, but not DMD MyoD transfected fibroblasts or skeletal muscle (Fig 6).

**Figure 6.**
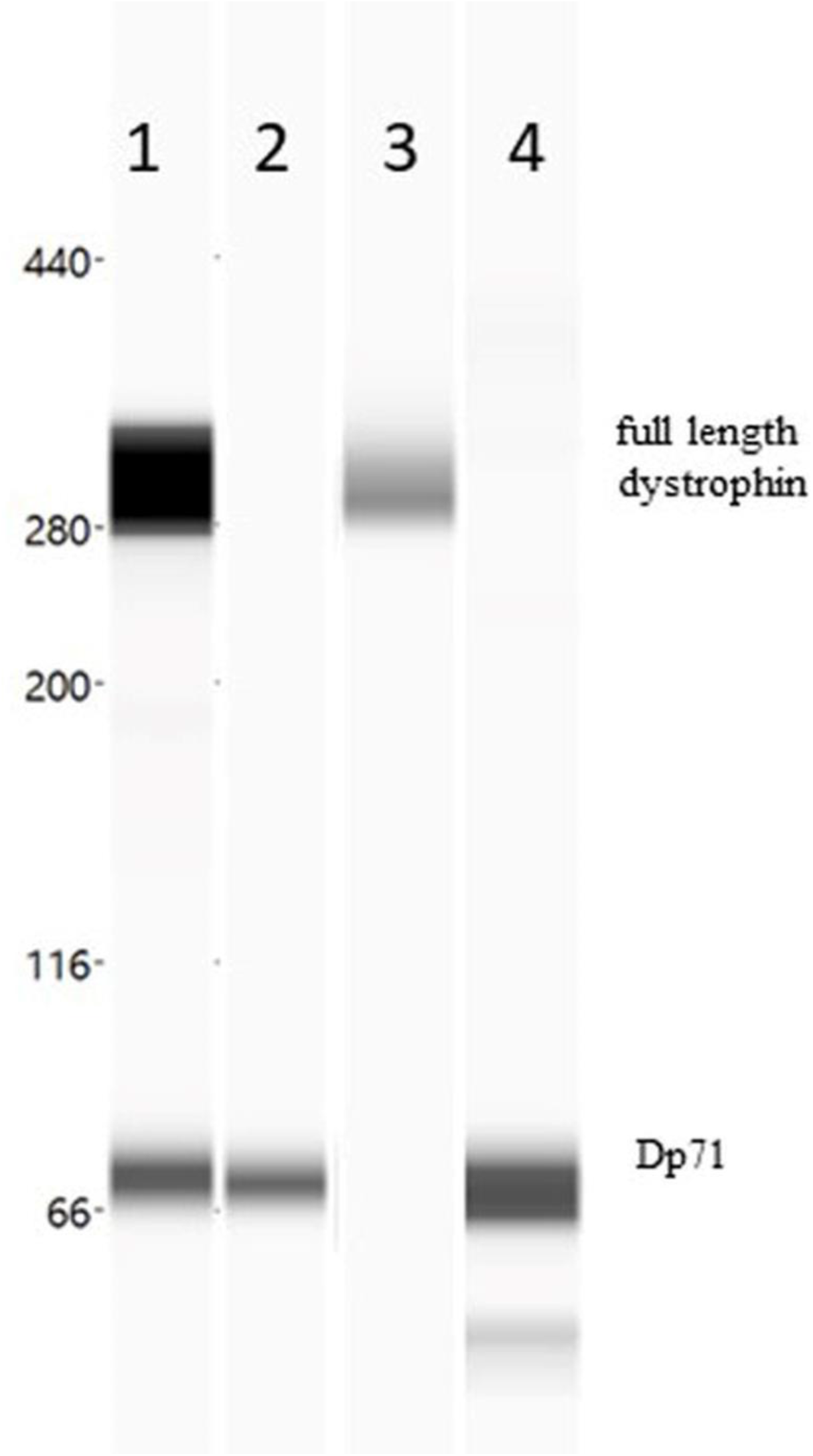
**Virtual blot of dystrophin signal detected by Wes using a C-terminus polyclonal anti dystrophin antibody (Abcam 15277) in: 1) control fibroblasts MyoD transfected; 2) DMD fibroblasts MyoD transfected; 3) control muscle; 4) DMD muscle. The amount of proteins used for each sample were: 1) 1µg; 2) 1µg; 3) 0.2501µg; 4) 0.1251µg. As previously reported, the full length dystrophin signal detected by Wes displays a molecular weight of approximately 300 kDa, which is lower than the predicted 427 kDa for the full-length (muscle) dystrophin isoform[25].**

## Discussion

DMD is characterised by significant variability in clinical progression[15,26]. Latent class trajectory analysis identified different classes of progression, but did not necessarily provide information on the drivers of the observed variability[15]. The reduced ability to identify drivers of this variability complicates assessments of DMD boys in clinical practice and clinical trials.

We hypothesised that *DMD* mutations expected to differentially impact on expression of different dystrophin isoforms could differentially affect motor function in DMD in both humans and mouse models. We focused on dystrophin isoforms previously demonstrated to play an important role in brain function. We performed a series of investigations characterising dystrophin isoform production in wild-type and DMD myogenic and skeletal muscle and characterising patterns of motor impairment in 3 DMD mouse models, wild-type mice and a large cohort of DMD boys with differential dystrophin isoform involvement.

Our results clearly demonstrate marked reductions in motor function achievement at 5 years of age (reduced mean NSAA scores and rise from supine times) and peak motor function (reduced mean peak NSAA scores and 10m walk/run velocities) in DMD boys lacking Dp140 and Dp71, with a clear cumulative effect of loss of isoforms. The differences in NSAA score observed between the isoform groups was considerable, substantially exceeding the minimally clinically important difference in NSAA score of approximately 3 points, highlighting the clear clinical significance of our findings[27].

We assessed the impact of loss of different dystrophin isoforms on grip strength in *mdx* (Dp427 absent, Dp140/Dp71 present); *mdx52* (Dp427/Dp140 absent, Dp71 present); and *DMD-null* mice (Dp427/Dp140/Dp71 absent). As expected, mdx mice had significantly reduced grip strength compared to wild type mice. When assessing the impact of isoform loss, average grip strength in *mdx* mice at 3 months of age was significantly higher than in *mdx52* mice (*p* = 0.01), but no further decline was observed in the *DMD-null* mice lacking all 3 isoforms.

When we studied dystrophin isoform protein production, we could not detect Dp140 in normal or DMD skeletal muscle or myogenic cultures. With the highly sensitive Wes technique, we detected low levels of Dp71 production in skeletal muscle of DMD boys, but not controls. This contrasted with the high Dp71 levels detected in both normal and DMD myogenic cultures.

This study directly correlates motor impairment in DMD humans and DMD mouse models to different patterns of dystrophin isoform expression.

A previous clinical study found DMD children with intellectual disability often have worse motor outcomes[28]. However, when genotype grouping was done, patients were subdivided between those with mutations proximal or distal to exon 30. Hence not allowing to distinguish the role of Dp140 and Dp71 in motor outcomes[28].

While our data unequivocally demonstrate differences in motor performance between these isoform groups, they do not necessarily provide a definitive mechanistic explanation for this. We considered a number of potential factors: differences in glucocorticoid use; a direct role of Dp140 and Dp71 in skeletal muscle function, differences in comprehension of motor outcome instructions; or impact of lack of Dp140 and Dp71 on higher aspects of motor coordination and planning.

With respect to glucocorticoid use, we found no difference in glucocorticoid regime between isoform groups and the pattern of cumulative marked reductions in motor outcomes in DMD boys lacking Dp140 and Dp71 remained present after adjusting for glucocorticoid regime.

Regarding the possibility that DMD boys lacking Dp140 and Dp71 were less able to understand instructions for performing assessments, we consider this unlikely as the components of the assessments are part of the children’s daily routine, the assessments are carried out every 6 months in hospital and the specialist physiotherapists have a standard protocol and regular training in carrying out the assessments.

With regards to the *mdx* mice study, the *mdx-52* mice had reduced grip strength compared to the *mdx* mice. However, we did not detect a significant difference in grip strength between the *DMD-null* and *mdx52* mice.

Our studies in DMD boys and dystrophic mouse models provide clear evidence of a relationship between patterns of DMD isoform involvement and motor performance and open the field to various possible explanations to account for these findings. The fact that the pattern of Dp140 expression is confined to the CNS makes the possibility of a skeletal muscle role for Dp140 not plausible. However, a possible, and previously not appreciated, role of Dp140 on motor neuron function cannot be excluded and could have contributed to the observed differences between Dp427 only and Dp427 and Dp140 deficient boys and mice.

Regarding Dp71, we confirm previous findings of Dp71 production in DMD muscle, but not control muscle[29]. In DMD muscle, Dp71 is not localised at the sarcolemma, C-terminal dystrophin staining is the standard diagnostic test and DMD patients universally demonstrate absent C-terminal dystrophin at the sarcolemma[30]. This observation, together with the absence of an actin binding domain in Dp71, argues against a peripheral effect of this isoform in functional motor outcomes, which is also confirmed by the dystrophic mouse studies. We confirm that myotubes in culture produce high levels of Dp71, and that this capacity is retained in DMD patients with mutations upstream of the Dp71 promoter. This, and previous findings, strongly, indicate a role for Dp71 in myoblast cell proliferation and satellite cell activation[31–33]. This suggests that the low level of Dp71 production in DMD muscle is a secondary phenomenon related to the muscle degeneration and regeneration characteristic of DMD.

One hypothesis we consider plausible is that the observed differences in motor function could be related to a direct impact of CNS involvement on motor function[11–13,28]. Whilst the concept of an impact of brain function on motor performance is novel in the DMD field, this is a well-recognised concept in other conditions. Westendorp *et al*. compared gross motor skills of children with learning disabilities (LD) with age-matched typically developing children[34]. Children with LD had lower locomotor scores than typically developing children[34]. Several studies have found that children with ASD without skeletal muscle involvement have both gross and fine motor delay[35,36]. These observations are relevant as the prevalence of LD is higher in DMD boys, particularly in those lacking Dp140 and Dp71[10].

Previous studies have hypothesised a possible role of the cerebellum and/or the cerebellar thalamic cortical connectivity in DMD cognitive difficulties[37]. Battini et al found a deficits of executive function, specifically in tasks requiring planning and directing goal oriented behaviour, in DMD boys[37]. Hellebrekers et al found Dp140 negative males had slower information processing speeds than Dp140 positive males[38].

With reference to the *mdx* mice, Chaussenot et al demonstrated executive dysfunction in a Dp71 deficient mouse model, whilst no deep CNS phenotyping is available for *mdx52* mice[39].

Dp427^m^, Dp140 and Dp71 are all expressed in the adult human cerebellum[7]. The cerebellum plays a crucial role in the control of goal-directed movements and timing of coordinated movement[40]. Features of cerebellar movement dysfunction in children include gross motor delay and poor coordination with complex movements (with relative sparing of simple motor tasks)[41]. Interestingly, of the 3 main ambulatory motor outcomes we considered, the NSAA score was the only outcome showing impairment in those lacking the shorter isoforms at both 5 years and peak motor function. Of these 3 outcomes, the NSAA is the most complex, requiring the highest level of motor coordination and planning.

Taken together, we consider a plausible contributor for the patterns of more severe motor impairment in boys and *mdx* mice lacking Dp140 and Dp140/Dp71 to be the associated deficits in the higher centres of motor function, control and coordination, with a possible role of deficits in the cerebellum and/or the cerebellar thalamic cortical connectivity.

The strengths of this study include the large number of patients, data collected in a ‘real world’ clinical setting and the longitudinal nature of the data. Limitations included some missing data due to the real-world data collection in routine clinical appointments. However, all missing data was accounted for statistically.

In summary, we found that mutations resulting in lack of Dp140 or Dp140/Dp71 in addition to Dp427 are associated with poorer motor function performance in DMD boys and dystrophic mouse models, with a cumulative effect of loss of isoforms in DMD boys. Given the major role of these isoforms in brain function, we hypothesise this could be at least partly related to deficits in the higher centres of motor function, control and coordination in those lacking Dp140 and Dp140/Dp71.

Irrespective of the possible pathophysiological explanation, our novel findings provide evidence for a relationship between the site of the DMD mutations and the effect on shorter dystrophin isoform production on not only cognitive, but also motor outcomes. This has crucial implications for clinical practice and clinical trial design involving patients with DMD.

## Supporting information

STROBE checklist

COI disclosures

COI disclosure

## Data Availability

The data that support the findings of this study are available from the corresponding author, [FM], upon reasonable request.

## Acknowledgements

We are grateful to the DMD patients and their families, the North Star clinical network study group and its senior clinico-academic coordinator Dr Vandana Ayyar Gupta, which is co-led by Francesco Muntoni and Adnan Y Manzur, Muscular Dystrophy UK (MDUK) for funding the North Star network and Certus Technology Associates Limited for hosting the database. Note that the lead authors for the North Star Group are Dr Francesco Muntoni; and Dr Adnan Manzur. The North Star DMD Network is supported by a grant from Muscular Dystrophy UK to Dr Adnan Manzur and Prof Francesco Muntoni, at UCL. We are very grateful to Cyrille Vaillend (Neuroscience Paris-Saclay Institute (Neuro-PSI), UMR 9197, Université Paris Sud, CNRS, Université Paris Saclay, 91190 Orsay, France) for his helpful review of and feedback on the manuscript.

The support of L’Association Française contre les Myopathies for the ‘Outcome measures in Duchenne Muscular Dystrophy: A Natural History Study: Imaging and Neuropsychology Sub-study’ is gratefully acknowledged. The support of Great Ormond Street Hospital Children’s Charity for ‘A Study of Emotional Function in Duchenne Muscular Dystrophy (EmoDe Study)’ is gratefully acknowledged. Kate Maresh was supported by the Medical Research Council (MRC) via the MRC Centre for Neuromuscular Diseases, Institute of Neurology, University College London.

The support of Great Ormond Street Hospital Children’s Charity and the NIHR GOSH biomedical research centre (BRC) for the ‘Neurodevelopmental, emotional, and behavioural problems in Duchenne muscular dystrophy in relation to underlying dystrophin gene mutations’ study is gratefully acknowledged^19^.

This work is supported by the NIHR GOSH BRC. The views expressed are participants of the author(s) and not necessarily participants of the NHS, the NIHR or the Department of Health.

The support of EUH2020 grant 83245 **B**rain **I**nvolvement i**N D**ystrophinopathies is also gratefully acknowledged.

The support of the grant Grants in Aid for Research on Nervous and Mental Disorders (grant number 2-6 to Yoshitsugu Aoki) for the functional testing using dystrophic and wild-type mice is gratefully acknowledged.

## Author’s financial relationships and potential conflicts of interest

- Mary Chesshyre, Deborah Ridout, Kate Maresh, Lianne Abbott, Vandana Ayyar Gupta, Marion Main, Yoko Ookubo, Yasumasa Hashimoto and Silvia Torelli report no financial disclosures or potential conflicts of interest.
- Yoshitsugu Aoki is the grant holder of the grant Grants in Aid for Research on Nervous and Mental Disorders (grant number 2-6)
- Kate Maresh has received payments to her institution from Great Ormond Street Hospital Children’s Charity and MRC Centre for Neuromuscular Diseases, Queen Square, London in the last 36 months.
- Valeria Ricotti – Co-founder, EVP and CMO of DiNAQOR, shareholder of Solid Biosciences
- Adnan Manzur is the clinical lead for the North Star clinical network and is one of the co-grant holders from the MDUK for maintenance of the network.
- Mariacristina Scoto has received speaker and consultancy honoraria For Roche, Avexis, Santhera and Biogen
- Giovanni Baranello has received consultancy honoraria from AveXis, Roche, Biogen, PTC, and Sarepta Therapeutics. Dr Giovanni Baranello has received speaker honoraria from AveXis, Roche and PTC
- Francesco Muntoni is supported by the NIHR Great Ormond Street Hospital Biomedical Research Centre and has received speaker and consultancy honoraria from Sarepta Therapeutics, Avexis, PTC Therapeutics, Roche and Pfizer.

## Ethical guidelines and consent

The North Star clinical network study was reviewed by the North Sheffield Research Ethics Committee and it was felt that it didn’t need to go to a research ethics committee. Written informed consent was obtained for the collection of all clinical data and the NorthStar clinical network project has Caldicott Guardian approval. All clinical assessments are conducted according to the principles of the Declaration of Helsinki (2000) and its later amendments and the Principles of Good Clinical Practice.

Animal studies were performed in accordance with the appropriate ethics committee and have therefore been performed with the ethical standards laid down in the 1964 Declaration of Helsinki and its later amendments.

## Notes

### Funding Statement

We are grateful to Muscular Dystrophy UK (MDUK) for funding the North Star network and Certus Technology Associates Limited for hosting the database. The North Star DMD Network is supported by a grant from Muscular Dystrophy UK to Dr Adnan Manzur and Prof Francesco Muntoni, at UCL.
The support of Association Francaise contre les Myopathies for the Outcome measures in Duchenne Muscular Dystrophy: A Natural History Study: Imaging and Neuropsychology Sub-study is gratefully acknowledged. The support of Great Ormond Street Hospital Childrens Charity for A Study of Emotional Function in Duchenne Muscular Dystrophy (EmoDe Study) is gratefully acknowledged. Kate Maresh was supported by the Medical Research Council (MRC) via the MRC Centre for Neuromuscular Diseases, Institute of Neurology, University College London.
The support of Great Ormond Street Hospital Childrens Charity and the NIHR GOSH biomedical research centre (BRC) for the Neurodevelopmental, emotional, and behavioural problems in Duchenne muscular dystrophy in relation to underlying dystrophin gene mutations study is gratefully acknowledged.
This work is supported by the NIHR GOSH BRC. The views expressed are participants of the author(s) and not necessarily participants of the NHS, the NIHR or the Department of Health.
The support of EUH2020 grant 83245 Brain Involvement iN Dystrophinopathies is also gratefully acknowledged.
The support of the grant Grants in Aid for Research on Nervous and Mental Disorders (grant number 2 to 6 to Yoshitsugu Aoki) for the functional testing using dystrophic and wild-type mice is gratefully acknowledged.

### Author Declarations

The North Star clinical network study was reviewed by the North Sheffield Research Ethics Committee and it was felt that it did not need to go to a research ethics committee. Written informed consent was obtained for the collection of all clinical data and the NorthStar clinical network project has Caldicott Guardian approval. All clinical assessments are conducted according to the principles of the Declaration of Helsinki (2000) and its later amendments and the Principles of Good Clinical Practice. Tissue sampling was approved by the NHS national Research Ethics Service as follows under the studies Setting up of a rare diseases biological samples bank (biobank) for research to facilitate pharmacological, gene and cell therapy trials in neuromuscular disorders (NMD) (REC reference number: 06/Q0406/33 - Hammersmith and Queen Charlottes and Chelsea Research Ethics Committee, The use of cells as a model system to study pathogenesis and therapeutic strategies for Neuromuscular Disorders (REC reference 13/LO/1826 - London - Stanmore Research Ethics Committee) and Genes and Proteins in Neuromuscular Disorders (REC reference 13/LO/1894 - London - Camberwell St Giles Research Ethics Committee). Animal studies were performed in accordance with the appropriate ethics committee and have therefore been performed with the ethical standards laid down in the 1964 Declaration of Helsinki and its later amendments.

## References

1. Darras BT, Jones HR, Ryan MM, De Vivo DC. Neuromuscular disorders of infancy, childhood, and adolescence : a clinician’s approach / edited by Basil T. Darras, MD, Joseph J. Volpe Professor of Neurology, Harvard Medical School, Associate Neurologist-in-Chief, Chief, Division of Clinical Neurology,. Second edi. London : Academic Press; 2015.

2. Mercuri E, Bönnemann CG, Muntoni F. Muscular dystrophies. Lancet. 2019 Nov;394(10213):2025–38.

3. Anthony K, Arechavala-Gomeza V, Ricotti V, Torelli S, Feng L, Janghra N, et al. Biochemical characterization of patients with in-frame or out-of-frame DMD deletions pertinent to exon 44 or 45 skipping. JAMA Neurol [Internet]. 2014 Jan 1 [cited 2018 Feb 25];71(1):32–40. Available from: http://archneur.jamanetwork.com/article.aspx?doi=10.1001/jamaneurol.2013.4908

4. Heydemann A, Ceco E, Lim JE, Hadhazy M, Ryder P, Moran JL, et al. Latent TGF-beta-binding protein 4 modifies muscular dystrophy in mice. J Clin Invest. 2009 Dec;119(12):3703–12.

5. Vetrone SA, Montecino-Rodriguez E, Kudryashova E, Kramerova I, Hoffman EP, Liu SD, et al. Osteopontin promotes fibrosis in dystrophic mouse muscle by modulating immune cell subsets and intramuscular TGF-beta. J Clin Invest [Internet]. 2009;119(6):1583–94. Available from: http://ovidsp.ovid.com/ovidweb.cgi?T=JS&PAGE=reference&D=med6&NEWS=N&AN=19451692

6. Bello L, Pegoraro E. The “Usual Suspects”: Genes for Inflammation, Fibrosis, Regeneration, and Muscle Strength Modify Duchenne Muscular Dystrophy. J Clin Med. 2019 May;8(5).

7. Doorenweerd N, Mahfouz A, van Putten M, Kaliyaperumal R, T’ Hoen PAC, Hendriksen JGM, et al. Timing and localization of human dystrophin isoform expression provide insights into the cognitive phenotype of Duchenne muscular dystrophy. Sci Rep. 2017 Oct;7(1):12575.

8. Muntoni F, Torelli S, Ferlini A. Dystrophin and mutations: one gene, several proteins, multiple phenotypes. Lancet Neurol. 2003 Dec;2(12):731–40.

9. Doorenweerd N, Straathof CS, Dumas EM, Spitali P, Ginjaar IB, Wokke BH, et al. Reduced cerebral gray matter and altered white matter in boys with Duchenne muscular dystrophy. Ann Neurol. 2014 Sep;76(3):403–11.

10. Ricotti V, Mandy WPL, Scoto M, Pane M, Deconinck N, Messina S, et al. Neurodevelopmental, emotional, and behavioural problems in Duchenne muscular dystrophy in relation to underlying dystrophin gene mutations. Dev Med Child Neurol. 2016 Jan;58(1):77–84.

11. Felisari G, Martinelli Boneschi F, Bardoni A, Sironi M, Comi GP, Robotti M, et al. Loss of Dp140 dystrophin isoform and intellectual impairment in Duchenne dystrophy. Neurology. 2000 Aug;55(4):559–64.

12. Daoud F, Angeard N, Demerre B, Martie I, Benyaou R, Leturcq F, et al. Analysis of Dp71 contribution in the severity of mental retardation through comparison of Duchenne and Becker patients differing by mutation consequences on Dp71 expression. Hum Mol Genet. 2009 Oct;18(20):3779–94.

13. Taylor PJ, Betts GA, Maroulis S, Gilissen C, Pedersen RL, Mowat DR, et al. Dystrophin gene mutation location and the risk of cognitive impairment in Duchenne muscular dystrophy. PLoS One. 2010 Jan;5(1):e8803.

14. Ricotti V, Ridout DA, Scott E, Quinlivan R, Robb SA, Manzur AY, et al. Longterm benefits and adverse effects of intermittent versus daily glucocorticoids in boys with Duchenne muscular dystrophy. J Neurol Neurosurg Psychiatry. 2013 Jun;84(6):698–705.

15. Muntoni F, Domingos J, Manzur AY, Mayhew A, Guglieri M, Sajeev G, et al. Categorising trajectories and individual item changes of the North Star Ambulatory Assessment in patients with Duchenne muscular dystrophy. PLoS One. 2019;14(9):e0221097.

16. Scott E, Eagle M, Mayhew A, Freeman J, Main M, Sheehan J, et al. Development of a functional assessment scale for ambulatory boys with Duchenne muscular dystrophy. Physiother Res Int. 2012 Jun;17(2):101–9.

17. Combs D, Edgin JO, Klewer S, Barber BJ, Morgan WJ, Hsu C-H, et al. OSA and Neurocognitive Impairment in Children With Congenital Heart Disease. Chest. 2020 Sep;158(3):1208–17.

18. Jenni OG, Fintelmann S, Caflisch J, Latal B, Rousson V, Chaouch A. Stability of cognitive performance in children with mild intellectual disability. Dev Med Child Neurol. 2015 May;57(5):463–9.

19. Bulfield G, Siller WG, Wight PA, Moore KJ. X chromosome-linked muscular dystrophy (mdx) in the mouse. Proc Natl Acad Sci U S A. 1984 Feb;81(4):1189–92.

20. D.S. A. Gene therapy for disorders affecting children, progress and potential. J Paediatr Child Health [Internet]. 2007;43(5):323–30. Available from: http://ovidsp.ovid.com/ovidweb.cgi?T=JS&PAGE=reference&D=emed11&NE WS=N&AN=46698296

21. Bulfield G, Siller WG, Wight PAL, Moore KJ. X chromosome-linked muscular dystrophy (mdx) in the mouse. Proc Natl Acad Sci U S A [Internet]. 1984 [cited 2021 Apr 11];81(4 I):1189–92. Available from: https://pubmed.ncbi.nlm.nih.gov/6583703/

22. Kudoh H, Ikeda H, Kakitani M, Ueda A, Hayasaka M, Tomizuka K, et al. A new model mouse for Duchenne muscular dystrophy produced by 2.4 Mb deletion of dystrophin gene using Cre-loxP recombination system. Biochem Biophys Res Commun. 2005 Mar;328(2):507–16.

23. Araki E, Nakamura K, Nakao K, Kameya S, Kobayashi O, Nonaka I, et al. Targeted disruption of exon 52 in the mouse dystrophin gene induced muscle degeneration similar to that observed in Duchenne muscular dystrophy. Biochem Biophys Res Commun. 1997 Sep;238(2):492–7.

24. Luca, Annamaria De (Sezione di Farmacologia, Dipartimento Farmacobiologico, Facoltà di Farmacia, Università di Bari I and the T-N working group. Use of grip strength meter to assess the limb strength of mdx mice. DMD_M.2.2.001 version 2.0 [Internet]. Treat NMD neuromuscular network. 2019 [cited 2021 Apr 11]. Available from: https://treat-nmd.org/wp-content/uploads/2016/08/MDX-DMD_M.2.2.001.pdf

25. Beekman C, Janson AA, Baghat A, van Deutekom JC, Datson NA. Use of capillary Western immunoassay (Wes) for quantification of dystrophin levels in skeletal muscle of healthy controls and individuals with Becker and Duchenne muscular dystrophy. PLoS One. 2018;13(4):e0195850.

26. Mercuri E, Signorovitch JE, Swallow E, Song J, Ward SJ, Group DMDI, et al. Categorizing natural history trajectories of ambulatory function measured by the 6-minute walk distance in patients with Duchenne muscular dystrophy. Pane M Messina S, Vita GL, Sormani MP, D’Amico A, Berardinelli A, Magri F, Comi GP, Baranello G, Mongini T, Pini A, Battini R, Pegoraro E, Bruno C, Politano L, Previtali S, Binks MH, Campion G, Charnas L, Kaye E, Kelly M, Morris C, Reha A ME, editor. Neuromuscul Disord [Internet]. 2016;26(9):576–83. Available from: http://ovidsp.ovid.com/ovidweb.cgi?T=JS&PAGE=reference&D=medc&NEWS=N&AN=27423700

27. Muntoni F, Manzur A MA et al. P.299 Minimal Detectable Change in the North Star Ambulatory assessment (NSAA) in Duchenne muscular dystrophy (DMD). Neuromuscul Disord. 2018;28(S121).

28. Desguerre I, Christov C, Mayer M, Zeller R, Becane H-M, Bastuji-Garin S, et al. Clinical heterogeneity of duchenne muscular dystrophy (DMD): definition of sub-phenotypes and predictive criteria by long-term follow-up. PLoS One. 2009;4(2):e4347.

29. Beekman C, Janson AA, Baghat A, van Deutekom JC, Datson NA. Use of capillary Western immunoassay (Wes) for quantification of dystrophin levels in skeletal muscle of healthy controls and individuals with Becker and Duchenne muscular dystrophy. PLoS One [Internet]. 2018 Apr 1 [cited 2021 Apr 11];13(4). Available from: https://pubmed.ncbi.nlm.nih.gov/29641567/

30. Muscle Biopsy - 5th Edition [Internet]. [cited 2021 Apr 11]. Available from: https://www.elsevier.com/books/muscle-biopsy/dubowitz/978-0-7020-7471-4

31. Farea M, Rani AQM, Maeta K, Nishio H, Matsuo M. Dystrophin Dp71ab is monoclonally expressed in human satellite cells and enhances proliferation of myoblast cells. Sci Rep [Internet]. 2020 Dec 1 [cited 2021 Apr 11];10(1). Available from: https://pubmed.ncbi.nlm.nih.gov/33051488/

32. Tennyson CN, Dally GY, Ray PN, Worton RG. Expression of the dystrophin isoform Dp71 in differentiating human fetal myogenic cultures. Hum Mol Genet. 1996 Oct;5(10):1559–66.

33. Howard PL, Dally GY, Ditta SD, Austin RC, Worton RG, Klamut HJ, et al. Dystrophin isoforms DP71 and DP427 have distinct roles in myogenic cells. Muscle Nerve. 1999 Jan;22(1):16–27.

34. Westendorp M, Hartman E, Houwen S, Smith J, Visscher C. The relationship between gross motor skills and academic achievement in children with learning disabilities. Res Dev Disabil [Internet]. 2011;32(6):2773–9. Available from: http://www.sciencedirect.com/science/article/pii/S0891422211002186

35. Lloyd M, MacDonald M, Lord C. Motor skills of toddlers with autism spectrum disorders. Autism [Internet]. 2011/05/24. 2013 Mar;17(2):133–46. Available from: https://pubmed.ncbi.nlm.nih.gov/21610184

36. Uljarevic M, Hedley D, Alvares GA, Varcin KJ, Whitehouse AJO. Relationship between early motor milestones and severity of restricted and repetitive behaviors in children and adolescents with autism spectrum disorder. Autism Res. 2017 Jun;10(6):1163–8.

37. Battini R, Chieffo D, Bulgheroni S, Piccini G, Pecini C, Lucibello S, et al. Cognitive profile in Duchenne muscular dystrophy boys without intellectual disability: The role of executive functions. Neuromuscul Disord. 2018 Feb;28(2):122–8.

38. Hellebrekers DMJ, Doorenweerd N, Sweere DJJ, van Kuijk SMJ, Aartsma-Rus AM, Klinkenberg S, et al. Longitudinal follow-up of verbal span and processing speed in Duchenne muscular dystrophy. Eur J Paediatr Neurol EJPN Off J Eur Paediatr Neurol Soc. 2020 Mar;25:120–6.

39. Chaussenot R, Amar M, Fossier P, Vaillend C. Dp71-Dystrophin Deficiency Alters Prefrontal Cortex Excitation-Inhibition Balance and Executive Functions. Mol Neurobiol. 2019 Apr;56(4):2670–84.

40. Manto M, Bower JM, Conforto AB, Delgado-García JM, da Guarda SNF, Gerwig M, et al. Consensus paper: roles of the cerebellum in motor control--the diversity of ideas on cerebellar involvement in movement. Cerebellum [Internet]. 2012 Jun;11(2):457–87. Available from: https://pubmed.ncbi.nlm.nih.gov/22161499

41. Salman MS, Tsai P. The Role of the Pediatric Cerebellum in Motor Functions, Cognition, and Behavior: A Clinical Perspective. Neuroimaging Clin N Am. 2016 Aug;26(3):317–29.

