## Supplementary data for "Duchenne muscular dystrophy patients lacking the dystrophin isoforms Dp140 and Dp71 and mouse models lacking Dp140 have a more severe motor phenotype"

**Supplementary table 1. Relationships between dystrophin isoform group and cognition and cognition group and peak NSAA scores**

| Relationship between dystrophin isoform group and cognition | | | |
|---|---|---|---|
| **Dystrophin isoform group** | **Normal cognition**<br>**No. (%)** | **Cognitive impairment**<br>**No. (%)** | |
| **Group 1 (n, %)** | 43 (68%) | 20 (32%) | |
| **Group 2 (n, %)** | 26 (47%) | 29 (53%) | |
| **Group 3 (n, %)** | 2 (22%) | 7 (78%) | |
| **Age of diagnosis in years**<br>**Mean (sd)** | 3.4 (1.7) | 3.1 (1.7) | |
| **Age of starting GC in years Mean (sd)** | 5.1 (1.4) | 5.2 (0.8) | |
| **Peak NSAA scores** | | | |
| | **Normal cognition**<br>**Mean (sd)** | **Impaired cognition**<br>**Mean (sd)** | **Both groups combined**<br>**Mean (sd)** |
| **Peak NSAA score overall (all GC regimens combined)** | 27.7<br>(5.3)<br>n=71 | 25.3<br>(5.9)<br>n=56 | 26.6<br>(5.7)<br>n=127 |

**Supplementary table 1. Relationships between dystrophin isoform group and cognition and cognition group and peak NSAA scores. GC=glucocorticoid. This table considers the subset of participants (n=127) for whom cognition grouping, GC regimen and peak NSAA scores were available.**

**Supplementary table 2. Additional members of the North Star clinical network**

| Name and highest degree | Location | Role in study | Contribution to study |
|---|---|---|---|
| Pinki Munot MD | Dubowitz Neuromuscular Centre, Great Ormond Street Hospital for Children NHS Trust | Site investigator | Data acquisition |
| Rosaline Quinlivan MD | Dubowitz Neuromuscular Centre, Great Ormond Street Hospital for Children NHS Trust, United Kingdom | Site investigator | Data acquisition |
| Anna Sarkozy MD PhD | Dubowitz Neuromuscular Centre, Great Ormond Street Hospital for Children NHS Trust, United Kingdom | Site investigator | Data acquisition |

| | | | |
|---|---|---|---|
| Volker Straub MD | Institute of Genetic Medicine, Newcastle, United Kingdom | Site investigator | Data acquisition |
| Michela Guglieri | Institute of Genetic Medicine, Newcastle, United Kingdom | Site investigator | Data acquisition |
| Anna Mayhew PhD | Institute of Genetic Medicine, Newcastle, United Kingdom | Site physiotherapist | Data acquisition |
| Meredith James PT | Institute of Genetic Medicine, Newcastle, United Kingdom | Site physiotherapist | Data acquisition |
| Robert Muni Lofra MSc | Institute of Genetic Medicine, Newcastle, United Kingdom | Site physiotherapist | Data acquisition |
| Dionne Moat BSc | Institute of Genetic Medicine, Newcastle, United Kingdom | Site physiotherapist | Data acquisition |
| Jassi Sodhi MSc | Institute of Genetic Medicine, Newcastle, United Kingdom | Site physiotherapist | Data acquisition |
| Zoya Alhaswani MD | Birmingham Heartlands Hospital, Birmingham, United Kingdom | Site investigator | Data acquisition |
| Deepak Parasumaran FRCPCH | Birmingham Heartlands Hospital, Birmingham, United Kingdom | Site investigator | Data acquisition |
| Heather McMurchie DipPhys | Birmingham Heartlands Hospital, Birmingham, United Kingdom | Site physiotherapist | Data acquisition |
| Anne-Marie Childs MBChB | Yorkshire Regional Muscle Clinic, Leeds General Infirmary, United Kingdom | Site investigator | Data acquisition |
| Karen Pysden MBChB | Yorkshire Regional Muscle Clinic, Leeds General Infirmary, United Kingdom | Site investigator | Data acquisition |
| Stefan Spinty MRCPCH MMedSc | Alder Hey Hospital, Royal Liverpool Children’s NHS Trust, United Kingdom | Site investigator | Data acquisition |
| Laura Cheshman BScPhys | Alder Hey Hospital, Royal Liverpool Children’s NHS Trust, United Kingdom | Site physiotherapist | Data acquisition |
| Alison Shillington DipPhys | Alder Hey Hospital, Royal Liverpool Children’s NHS Trust, United Kingdom | Site physiotherapist | Data acquisition |
| Elizabeth Wraige MBBS BSc | Evelina London Children’s Hospital, Guy’s and St Thomas’ NHS Foundation Trust | Site investigator | Data acquisition |
| Heinz Jungbluth MD PhD | Evelina London Children’s Hospital, Guy’s and St Thomas’ NHS Foundation Trust | Site investigator | Data acquisition |

| | | | |
|---|---|---|---|
| Vasantha Gowda MBBS MSc | Evelina London Children's Hospital, Guy's and St Thomas' NHS Foundation Trust | Site investigator | Data acquisition |
| Jennie Sheehan MC SP | Evelina London Children's Hospital, Guy's and St Thomas' NHS Foundation Trust | Site physiotherapist | Data acquisition |
| Imelda Hughes MBBCh | Royal Manchester Children's Hospital, Manchester, United Kingdom | Site investigator | Data acquisition |
| Gary McCullagh MBBCh BAO MRCPCH | Royal Manchester Children's Hospital, Manchester, United Kingdom | Site investigator | Data acquisition |
| Sinead Warner BSc | Royal Manchester Children's Hospital, Manchester, United Kingdom | Site physiotherapist | Data acquisition |
| Tracey Willis MD | The Muscle Clinic, Robert Jones and Agnes Hunt Orthopaedic Hospital NHS Foundation Trust, United Kingdom | Site investigator | Data acquisition |
| Richa Kulshrestha MRCPCH | The Muscle Clinic, Robert Jones and Agnes Hunt Orthopaedic Hospital NHS Foundation Trust, United Kingdom | Site investigator | Data acquisition |
| Nicholas Emery BSc | The Muscle Clinic, Robert Jones and Agnes Hunt Orthopaedic Hospital NHS Foundation Trust, United Kingdom | Site physiotherapist | Data acquisition |
| Min Ong MRCPCH | Sheffield Children's Hospital NHS Foundation Trust, United Kingdom | Site investigator | Data acquisition |
| Frances Gibbon MB ChB | Department of Paediatric Neurology, University Hospital of Wales | Site investigator | Data acquisition |
| Bethan Parsons BSc | Department of Paediatric Neurology, University Hospital of Wales | Site physiotherapist | Data acquisition |
| Anirban Majumdar MRCPCH | Bristol Royal Hospital for Children, University Hospitals Bristol NHS Foundation Trust, United Kingdom | Site investigator | Data acquisition |
| Kayal Vijayakumar MRCPCH | Bristol Royal Hospital for Children, University Hospitals Bristol NHS Foundation Trust, United Kingdom | Site investigator | Data acquisition |
| Karen Naismith FRCPCH | Armistead Child Development Centre, Kings Cross Hospital, Scotland, United Kingdom | Site investigator | Data acquisition |

| | | | |
|---|---|---|---|
| Julie Burslem BSc | Armistead Child Development Centre, Kings Cross Hospital, Scotland, United Kingdom | Site physiotherapist | Data acquisition |
| Iain Horrocks MRCPCH | Fraser of Allander Neurosciences Unit, Royal Hospital for Sick Children, Glasgow, Scotland, United Kingdom | Site investigator | Data acquisition |
| Marina Di Marco MSc | Fraser of Allander Neurosciences Unit, Royal Hospital for Sick Children, Glasgow, Scotland, United Kingdom | Site physiotherapist | Data acquisition |
| Gabby Chow MD | Queen's Medical Centre, University Hospital, Nottingham, United Kingdom | Site investigator | Data acquisition |
| Christian deGoede MRCPCH | Preston Royal Hospital, Lancashire Teaching Hospitals NHS Foundation Trust, United Kingdom | Site investigator | Data acquisition |
| Andrea Selley MSc | Preston Royal Hospital, Lancashire Teaching Hospitals NHS Foundation Trust, United Kingdom | Site physiotherapist | Data acquisition |
| Neil Thomas FRCPCH | Southampton Children's Hospital, University Hospital Southampton NHS Foundation Trust, United Kingdom | Site investigator | Data acquisition |
| Marjorie Illingworth MBChB | Southampton Children's Hospital, University Hospital Southampton NHS Foundation Trust, United Kingdom | Site investigator | Data acquisition |
| Michelle Geary BScPhys | Southampton Children's Hospital, University Hospital Southampton NHS Foundation Trust, United Kingdom | Site investigator | Data acquisition |
| Jenni Palmer BScPhys | Southampton Children's Hospital, University Hospital Southampton NHS Foundation Trust, United Kingdom | Site investigator | Data acquisition |
| Cathy White MD | Morriston Hospital, Heol Maes Eglwys, Wales, United Kingdom | Site investigator | Data acquisition |
| Kate Greenfield BSc | Morriston Hospital, Heol Maes Eglwys, Wales, United Kingdom | Site physiotherapist | Data acquisition |
| Sandya Tirupathi MRCPCH | Royal Belfast Hospital for Sick Children, Belfast, Northern Ireland, United Kingdom | Site investigator | Data acquisition |

| | | | |
|---|---|---|---|
| Melanie Douglas BSc | Royal Belfast Hospital for Sick Children, Belfast, Northern Ireland, United Kingdom | Site physiotherapist | Data acquisition |
| Jaci McFetridge BSc | Royal Belfast Hospital for Sick Children, Belfast, Northern Ireland, United Kingdom | Site physiotherapist | Data acquisition |
| Gemunu Hewawitharana MBBS | Department of Paediatric Neurology, Leicester Royal Infirmary, United Kingdom | Site investigator | Data acquisition |
| Gautam Ambegaonkar MRCPCH DCH | Child Development Centre, Addenbrooke's Hospital, Cambridge University Hospitals NHS Foundation Trust, United Kingdom | Site investigator | Data acquisition |
| Deepa Krishnakumar MRCPCH DCH | Child Development Centre, Addenbrooke's Hospital, Cambridge University Hospitals NHS Foundation Trust, United Kingdom | Site investigator | Data acquisition |
| Catherine Ward BSc | Child Development Centre, Addenbrooke's Hospital, Cambridge University Hospitals NHS Foundation Trust, United Kingdom | Site physiotherapist | Data acquisition |
| Jacqui Taylor BSc | Child Development Centre, Addenbrooke's Hospital, Cambridge University Hospitals NHS Foundation Trust, United Kingdom | Site physiotherapist | Data acquisition |
| Elma Stephens MD | Royal Aberdeen Children's Hospital, Scotland, United Kingdom | Site investigator | Data acquisition |
| Jane Tewnion BSc | Royal Aberdeen Children's Hospital, Scotland, United Kingdom | Site physiotherapist | Data acquisition |
| Sithara Ramdas FRCPCH | Oxford Children's Hospital, John Radcliffe Hospital, United Kingdom | Site investigator | Data acquisition |
| Alex Baxter MRCPCH | Royal Hospital for Sick Children, Edinburgh, Scotland, United Kingdom | Site investigator | Data acquisition |

**Supplementary table 2. Additional members of the NorthStar clinical network - name and highest degree, location, role in study and contribution to study.**
