## Supplementary material for "Duchenne muscular dystrophy patients lacking the dystrophin isoforms Dp140 and Dp71 and mouse models lacking Dp140 have a more severe motor phenotype": COI disclosures

**ICMJE DISCLOSURE FORM**

**Date:________________10th July 2021**
**Your Name:______________________Francesco Muntoni**
**Manuscript Title:** Investigating the Role of Dystrophin Isoform Deficiency in Motor Function in Duchenne Muscular Dystrophy
**Manuscript number (if known):_______________________________________________________________________**

**In the interest of transparency, we ask you to disclose all relationships/activities/interests listed below that are related to the content of your manuscript. "Related" means any relation with for-profit or not-for-profit third parties whose interests may be affected by the content of the manuscript. Disclosure represents a commitment to transparency and does not necessarily indicate a bias. If you are in doubt about whether to list a relationship/activity/interest, it is preferable that you do so.**

**The following questions apply to the author's relationships/activities/interests as they relate to the current manuscript only.**

**The author's relationships/activities/interests should be defined broadly. For example, if your manuscript pertains to the epidemiology of hypertension, you should declare all relationships with manufacturers of antihypertensive medication, even if that medication is not mentioned in the manuscript.**

**In item #1 below, report all support for the work reported in this manuscript without time limit. For all other items, the time frame for disclosure is the past 36 months.**

| | | **Name all entities with whom you have this relationship or indicate none (add rows as needed)** | **Specifications/Comments (e.g., if payments were made to you or to your institution)** |
|---|---|---|---|
| | | **Time frame: Since the initial planning of the work** | |
| 1 | All support for the present manuscript (e.g., funding, provision of study materials, medical writing, article processing charges, etc.) **No time limit for this item.** | NIHR | Contribution to UCL for my work on Novel Therapies in the Biomedical Research Centre |
| | | European Commission grant EUH2020 83245 **B**rain **I**nvolvement i**N** **D**ystrophinopathies | Payment to UCL |
| | | **Time frame: past 36 months** | |
| 2 | Grants or contracts from any entity (if not indicated in item #1 above). | ____ None | |

| | | |
|---|---|---|
| 3 | Royalties or licenses | _____ None |
| 4 | Consulting fees | _____ None |
| 5 | Payment or honoraria for lectures, presentations, speakers bureaus, manuscript writing or educational events | _____ None |
| 6 | Payment for expert testimony | _____ None |
| 7 | Support for attending meetings and/or travel | _____ None |
| 8 | Patents planned, issued or pending | _____ None |
| 9 | Participation on a Data Safety Monitoring Board or Advisory Board | _____ None |
| 10 | Leadership or fiduciary role in other board, society, committee or advocacy group, paid or unpaid | _____ None |
| 11 | Stock or stock options | _____ None |
| 12 | Receipt of equipment, materials, drugs, medical writing, gifts or other services | _____ None |
| 13 | Other financial or non-financial interests | _____ None |

**Please place an "X" next to the following statement to indicate your agreement:**

**_x__ I certify that I have answered every question and have not altered the wording of any of the questions on this form.**

**Francesco Muntoni**

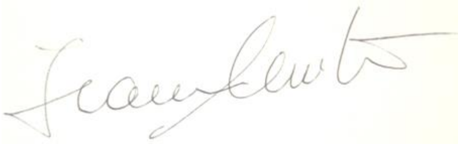

### ICMJE DISCLOSURE FORM

**Date: 13/7/2021**______________________________________________
**Your Name: Lianne Abbott** ______________________________________________
**Manuscript Title:** Investigating the Role of Dystrophin Isoform Deficiency in Motor Function in Duchenne Muscular Dystrophy
**Manuscript number (if known):**______________________________________________

**In the interest of transparency, we ask you to disclose all relationships/activities/interests listed below that are related to the content of your manuscript. "Related" means any relation with for-profit or not-for-profit third parties whose interests may be affected by the content of the manuscript. Disclosure represents a commitment to transparency and does not necessarily indicate a bias. If you are in doubt about whether to list a relationship/activity/interest, it is preferable that you do so.**

**The following questions apply to the author's relationships/activities/interests as they relate to the current manuscript only.**

**The author's relationships/activities/interests should be defined broadly. For example, if your manuscript pertains to the epidemiology of hypertension, you should declare all relationships with manufacturers of antihypertensive medication, even if that medication is not mentioned in the manuscript.**

**In item #1 below, report all support for the work reported in this manuscript without time limit. For all other items, the time frame for disclosure is the past 36 months.**

| | | **Name all entities with whom you have this relationship or indicate none (add rows as needed)** | **Specifications/Comments (e.g., if payments were made to you or to your institution)** |
|---|---|---|---|
| | | **Time frame: Since the initial planning of the work** | |
| 1 | All support for the present manuscript (e.g., funding, provision of study materials, medical writing, article processing charges, etc.) **No time limit for this item.** | ____ None | |
| | | **Time frame: past 36 months** | |
| 2 | Grants or contracts from any entity (if not indicated in item #1 above). | ____ None | |
| 3 | Royalties or licenses | ____ None | |

| | | |
|---|---|---|
| 4 | Consulting fees | ____ None |
| 5 | Payment or honoraria for lectures, presentations, speakers bureaus, manuscript writing or educational events | ____ None |
| 6 | Payment for expert testimony | ____ None |
| 7 | Support for attending meetings and/or travel | ____ None |
| 8 | Patents planned, issued or pending | ____ None |
| 9 | Participation on a Data Safety Monitoring Board or Advisory Board | ____ None |
| 10 | Leadership or fiduciary role in other board, society, committee or advocacy group, paid or unpaid | ____ None |
| 11 | Stock or stock options | ____ None |
| 12 | Receipt of equipment, materials, drugs, medical writing, gifts or other services | ____ None |
| 13 | Other financial or non-financial interests | ____ None |

**Please place an "X" next to the following statement to indicate your agreement:**

**_X_ I certify that I have answered every question and have not altered the wording of any of the questions on this form.**

**ICMJE DISCLOSURE FORM**

**Date:** 15 July 2021
**Your Name:** Adnan Manzur
**Manuscript Title:** Investigating the Role of Dystrophin Isoform Deficiency in Motor Function in Duchenne Muscular Dystrophy
**Manuscript number (if known):**_______________________________________________________________

**In the interest of transparency, we ask you to disclose all relationships/activities/interests listed below that are related to the content of your manuscript. "Related" means any relation with for-profit or not-for-profit third parties whose interests may be affected by the content of the manuscript. Disclosure represents a commitment to transparency and does not necessarily indicate a bias. If you are in doubt about whether to list a relationship/activity/interest, it is preferable that you do so.**

**The following questions apply to the author's relationships/activities/interests as they relate to the current manuscript only.**

**The author's relationships/activities/interests should be defined broadly. For example, if your manuscript pertains to the epidemiology of hypertension, you should declare all relationships with manufacturers of antihypertensive medication, even if that medication is not mentioned in the manuscript.**

**In item #1 below, report all support for the work reported in this manuscript without time limit. For all other items, the time frame for disclosure is the past 36 months.**

| | | **Name all entities with whom you have this relationship or indicate none (add rows as needed)** | **Specifications/Comments (e.g., if payments were made to you or to your institution)** |
|---|---|---|---|
| | | **Time frame: Since the initial planning of the work** | |
| 1 | All support for the present manuscript (e.g., funding, provision of study materials, medical writing, article processing charges, etc.) **No time limit for this item.** | ___Yes | Dr Adnan Manzur is one of the co- grant holders from the MDUK for maintenance of the NorthStar network. |
| | | **Time frame: past 36 months** | |
| 2 | Grants or contracts from any entity (if not indicated in item #1 above). | ____X None | |

| | | |
|---|---|---|
| 3 | Royalties or licenses | _____ X None |
| 4 | Consulting fees | _____X None |
| 5 | Payment or honoraria for lectures, presentations, speakers bureaus, manuscript writing or educational events | _____ X None |
| 6 | Payment for expert testimony | _____X None |
| 7 | Support for attending meetings and/or travel | _____X None |
| 8 | Patents planned, issued or pending | _____ X None |
| 9 | Participation on a Data Safety Monitoring Board or Advisory Board | _____ X None |
| 10 | Leadership or fiduciary role in other board, society, committee or advocacy group, paid or unpaid | _____ X None |
| 11 | Stock or stock options | _____ X None |
| 12 | Receipt of equipment, materials, drugs, medical writing, gifts or other services | _____ X None |
| 13 | Other financial or non-financial interests | _____ X None |

**Please place an "X" next to the following statement to indicate your agreement:**

**_X I certify that I have answered every question and have not altered the wording of any of the questions on this form.**

**ICMJE DISCLOSURE FORM**

**Date:** 15 July 2021
**Your Name:** Giovanni Baranello
**Manuscript Title:** Investigating the Role of Dystrophin Isoform Deficiency in Motor Function in Duchenne Muscular Dystrophy
**Manuscript number (if known):**_______________________________________________________________

**In the interest of transparency, we ask you to disclose all relationships/activities/interests listed below that are related to the content of your manuscript. "Related" means any relation with for-profit or not-for-profit third parties whose interests may be affected by the content of the manuscript. Disclosure represents a commitment to transparency and does not necessarily indicate a bias. If you are in doubt about whether to list a relationship/activity/interest, it is preferable that you do so.**

**The following questions apply to the author's relationships/activities/interests as they relate to the current manuscript only.**

**The author's relationships/activities/interests should be defined broadly. For example, if your manuscript pertains to the epidemiology of hypertension, you should declare all relationships with manufacturers of antihypertensive medication, even if that medication is not mentioned in the manuscript.**

**In item #1 below, report all support for the work reported in this manuscript without time limit. For all other items, the time frame for disclosure is the past 36 months.**

| | | Name all entities with whom you have this relationship or indicate none (add rows as needed) | Specifications/Comments (e.g., if payments were made to you or to your institution) |
|---|---|---|---|
| **Time frame: Since the initial planning of the work** | | | |
| 1 | All support for the present manuscript (e.g., funding, provision of study materials, medical writing, article processing charges, etc.) **No time limit for this item.** | ___X None | |
| **Time frame: past 36 months** | | | |
| 2 | Grants or contracts from any entity (if not indicated in item #1 above). | ____X None | |
| 3 | Royalties or licenses | ____ X None | |

| | | | |
|---|---|---|---|
| 4 | Consulting fees | _____Yes | Dr Giovanni Baranello has received consultancy honoraria from AveXis, Roche, Biogen, PTC, and Sarepta Therapeutics |
| 5 | Payment or honoraria for lectures, presentations, speakers bureaus, manuscript writing or educational events | _____ Yes | Dr Giovanni Baranello has received speaker honoraria from AveXis, Roche and PTC |
| 6 | Payment for expert testimony | _____X None | |
| 7 | Support for attending meetings and/or travel | _____X None | |
| 8 | Patents planned, issued or pending | _____ X None | |
| 9 | Participation on a Data Safety Monitoring Board or Advisory Board | _____ X None | |
| 10 | Leadership or fiduciary role in other board, society, committee or advocacy group, paid or unpaid | _____ X None | |
| 11 | Stock or stock options | _____ X None | |
| 12 | Receipt of equipment, materials, drugs, medical writing, gifts or other services | _____ | Dr Giovanni Baranello has received support from Roche for purchasing equipment to improve patients' nutritional management at his institution |
| 13 | Other financial or non-financial interests | _____ X None | |

**Please place an "X" next to the following statement to indicate your agreement:**

**___ I certify that I have answered every question and have not altered the wording of any of the questions on this form.**

**ICMJE DISCLOSURE FORM**

**Date:** 26 July 2021
**Your Name:** Mary Chesshyre
**Manuscript Title:** Duchenne muscular dystrophy patients lacking the dystrophin isoforms Dp140 and Dp71 and mouse models lacking Dp140 have a more severe motor phenotype

**Manuscript number (if known):**________________________________________________________________

**In the interest of transparency, we ask you to disclose all relationships/activities/interests listed below that are related to the content of your manuscript. "Related" means any relation with for-profit or not-for-profit third parties whose interests may be affected by the content of the manuscript. Disclosure represents a commitment to transparency and does not necessarily indicate a bias. If you are in doubt about whether to list a relationship/activity/interest, it is preferable that you do so.**

**The following questions apply to the author's relationships/activities/interests as they relate to the current manuscript only.**

**The author's relationships/activities/interests should be defined broadly. For example, if your manuscript pertains to the epidemiology of hypertension, you should declare all relationships with manufacturers of antihypertensive medication, even if that medication is not mentioned in the manuscript.**

**In item #1 below, report all support for the work reported in this manuscript without time limit. For all other items, the time frame for disclosure is the past 36 months.**

| | | **Name all entities with whom you have this relationship or indicate none (add rows as needed)** | **Specifications/Comments (e.g., if payments were made to you or to your institution)** |
|---|---|---|---|
| **Time frame: Since the initial planning of the work** | | | |
| 1 | All support for the present manuscript (e.g., funding, provision of study materials, medical writing, article processing charges, etc.) **No time limit for this item.** | ____X None | |
| **Time frame: past 36 months** | | | |
| 2 | Grants or contracts from any entity (if not indicated in item #1 above). | __X__ None | |

| | | |
|---|---|---|
| 3 | Royalties or licenses | __X__ None |
| 4 | Consulting fees | __X__ None |
| 5 | Payment or honoraria for lectures, presentations, speakers bureaus, manuscript writing or educational events | __X__ None |
| 6 | Payment for expert testimony | __X__ None |
| 7 | Support for attending meetings and/or travel | __X__ None |
| 8 | Patents planned, issued or pending | __X__ None |
| 9 | Participation on a Data Safety Monitoring Board or Advisory Board | __X__ None |
| 10 | Leadership or fiduciary role in other board, society, committee or advocacy group, paid or unpaid | __X__ None |
| 11 | Stock or stock options | __X__ None |
| 12 | Receipt of equipment, materials, drugs, medical writing, gifts or other services | __X__ None |
| 13 | Other financial or non-financial interests | __X__ None |

**Please place an "X" next to the following statement to indicate your agreement:**

**_X__ I certify that I have answered every question and have not altered the wording of any of the questions on this form.**

### ICMJE DISCLOSURE FORM

**Date:** 15 July 2021
**Your Name:** Marion Main
**Manuscript Title:** Investigating the Role of Dystrophin Isoform Deficiency in Motor Function in Duchenne Muscular Dystrophy
**Manuscript number (if known):**________________________________________________________________

**In the interest of transparency, we ask you to disclose all relationships/activities/interests listed below that are related to the content of your manuscript. "Related" means any relation with for-profit or not-for-profit third parties whose interests may be affected by the content of the manuscript. Disclosure represents a commitment to transparency and does not necessarily indicate a bias. If you are in doubt about whether to list a relationship/activity/interest, it is preferable that you do so.**

**The following questions apply to the author's relationships/activities/interests as they relate to the current manuscript only.**

**The author's relationships/activities/interests should be defined broadly. For example, if your manuscript pertains to the epidemiology of hypertension, you should declare all relationships with manufacturers of antihypertensive medication, even if that medication is not mentioned in the manuscript.**

**In item #1 below, report all support for the work reported in this manuscript without time limit. For all other items, the time frame for disclosure is the past 36 months.**

| | | **Name all entities with whom you have this relationship or indicate none (add rows as needed)** | **Specifications/Comments (e.g., if payments were made to you or to your institution)** |
|---|---|---|---|
| **Time frame: Since the initial planning of the work** | | | |
| 1 | All support for the present manuscript (e.g., funding, provision of study materials, medical writing, article processing charges, etc.) **No time limit for this item.** | X None | |
| **Time frame: past 36 months** | | | |
| 2 | Grants or contracts from any entity (if not indicated in item #1 above). | X None | |
| 3 | Royalties or licenses | ____ X None | |

| | | |
|---|---|---|
| 4 | Consulting fees | X None |
| 5 | Payment or honoraria for lectures, presentations, speakers bureaus, manuscript writing or educational events | X None |
| 6 | Payment for expert testimony | X None |
| 7 | Support for attending meetings and/or travel | X None |
| 8 | Patents planned, issued or pending | X None |
| 9 | Participation on a Data Safety Monitoring Board or Advisory Board | X None |
| 10 | Leadership or fiduciary role in other board, society, committee or advocacy group, paid or unpaid | X None |
| 11 | Stock or stock options | X None |
| 12 | Receipt of equipment, materials, drugs, medical writing, gifts or other services | X None |
| 13 | Other financial or non-financial interests | X None |

**Please place an “X” next to the following statement to indicate your agreement:**

**X I certify that I have answered every question and have not altered the wording of any of the questions on this form.**

**ICMJE DISCLOSURE FORM**

**Date:** 15 July 2021
**Your Name:** Mariacristina Scoto
**Manuscript Title:** Investigating the Role of Dystrophin Isoform Deficiency in Motor Function in Duchenne Muscular Dystrophy
**Manuscript number (if known):**_________________________________________________________________

**In the interest of transparency, we ask you to disclose all relationships/activities/interests listed below that are related to the content of your manuscript. "Related" means any relation with for-profit or not-for-profit third parties whose interests may be affected by the content of the manuscript. Disclosure represents a commitment to transparency and does not necessarily indicate a bias. If you are in doubt about whether to list a relationship/activity/interest, it is preferable that you do so.**

**The following questions apply to the author's relationships/activities/interests as they relate to the current manuscript only.**

**The author's relationships/activities/interests should be defined broadly. For example, if your manuscript pertains to the epidemiology of hypertension, you should declare all relationships with manufacturers of antihypertensive medication, even if that medication is not mentioned in the manuscript.**

**In item #1 below, report all support for the work reported in this manuscript without time limit. For all other items, the time frame for disclosure is the past 36 months.**

| | | Name all entities with whom you have this relationship or indicate none (add rows as needed) | Specifications/Comments (e.g., if payments were made to you or to your institution) |
|---|---|---|---|
| **Time frame: Since the initial planning of the work** | | | |
| 1 | All support for the present manuscript (e.g., funding, provision of study materials, medical writing, article processing charges, etc.) **No time limit for this item.** | ___X None | |
| **Time frame: past 36 months** | | | |
| 2 | Grants or contracts from any entity (if not indicated in item #1 above). | ____X None | |
| 3 | Royalties or licenses | ____ X None | |

| | | | |
|---|---|---|---|
| 4 | Consulting fees | ____Yes | Dr Mariacristina Scoto has received consultancy honoraria For Roche, Avexis, Santhera and Biogen |
| 5 | Payment or honoraria for lectures, presentations, speakers bureaus, manuscript writing or educational events | ____ Yes | Dr Mariacristina Scoto has received speaker honoraria For Roche, Avexis, Santhera and Biogen |
| 6 | Payment for expert testimony | ____X None | |
| 7 | Support for attending meetings and/or travel | ____X None | |
| 8 | Patents planned, issued or pending | ____ X None | |
| 9 | Participation on a Data Safety Monitoring Board or Advisory Board | ____ X None | |
| 10 | Leadership or fiduciary role in other board, society, committee or advocacy group, paid or unpaid | ____ X None | |
| 11 | Stock or stock options | ____ X None | |
| 12 | Receipt of equipment, materials, drugs, medical writing, gifts or other services | ____ X None | |
| 13 | Other financial or non-financial interests | ____ X None | |

**Please place an "X" next to the following statement to indicate your agreement:**

**X I (Mariacristina Scoto) certify that I have answered every question and have not altered the wording of any of the questions on this form.**

### ICMJE DISCLOSURE FORM

**Date:______12th July 2021________________________________________________________________________**
**Your Name:_____Ookubo Yoko__________________________________________________________________**
**Manuscript Title:** Investigating the Role of Dystrophin Isoform Deficiency in Motor Function in Duchenne Muscular Dystrophy
**Manuscript number (if known):__________________________________________________________________**

**In the interest of transparency, we ask you to disclose all relationships/activities/interests listed below that are related to the content of your manuscript. "Related" means any relation with for-profit or not-for-profit third parties whose interests may be affected by the content of the manuscript. Disclosure represents a commitment to transparency and does not necessarily indicate a bias. If you are in doubt about whether to list a relationship/activity/interest, it is preferable that you do so.**

**The following questions apply to the author's relationships/activities/interests as they relate to the current manuscript only.**

**The author's relationships/activities/interests should be defined broadly. For example, if your manuscript pertains to the epidemiology of hypertension, you should declare all relationships with manufacturers of antihypertensive medication, even if that medication is not mentioned in the manuscript.**

**In item #1 below, report all support for the work reported in this manuscript without time limit. For all other items, the time frame for disclosure is the past 36 months.**

| | | **Name all entities with whom you have this relationship or indicate none (add rows as needed)** | **Specifications/Comments (e.g., if payments were made to you or to your institution)** |
|---|---|---|---|
| | | **Time frame: Since the initial planning of the work** | |
| 1 | All support for the present manuscript (e.g., funding, provision of study materials, medical writing, article processing charges, etc.) **No time limit for this item.** | ____ None | |
| | | **Time frame: past 36 months** | |
| 2 | Grants or contracts from any entity (if not indicated in item #1 above). | ____ None | |
| 3 | Royalties or licenses | ____ None | |

| | | |
|---|---|---|
| 4 | Consulting fees | ____ None |
| 5 | Payment or honoraria for lectures, presentations, speakers bureaus, manuscript writing or educational events | ____ None |
| 6 | Payment for expert testimony | ____ None |
| 7 | Support for attending meetings and/or travel | ____ None |
| 8 | Patents planned, issued or pending | ____ None |
| 9 | Participation on a Data Safety Monitoring Board or Advisory Board | ____ None |
| 10 | Leadership or fiduciary role in other board, society, committee or advocacy group, paid or unpaid | ____ None |
| 11 | Stock or stock options | ____ None |
| 12 | Receipt of equipment, materials, drugs, medical writing, gifts or other services | ____ None |
| 13 | Other financial or non-financial interests | ____ None |

**Please place an "X" next to the following statement to indicate your agreement:**

**_X_ I certify that I have answered every question and have not altered the wording of any of the questions on this form.**

**ICMJE DISCLOSURE FORM**

**Date:** 15 July 2021
**Your Name:** Vandana Ayyar Gupta
**Manuscript Title:** Investigating the Role of Dystrophin Isoform Deficiency in Motor Function in Duchenne Muscular Dystrophy
**Manuscript number (if known):**_______________________________________________________________

**In the interest of transparency, we ask you to disclose all relationships/activities/interests listed below that are related to the content of your manuscript. "Related" means any relation with for-profit or not-for-profit third parties whose interests may be affected by the content of the manuscript. Disclosure represents a commitment to transparency and does not necessarily indicate a bias. If you are in doubt about whether to list a relationship/activity/interest, it is preferable that you do so.**

**The following questions apply to the author's relationships/activities/interests as they relate to the current manuscript only.**

**The author's relationships/activities/interests should be defined broadly. For example, if your manuscript pertains to the epidemiology of hypertension, you should declare all relationships with manufacturers of antihypertensive medication, even if that medication is not mentioned in the manuscript.**

**In item #1 below, report all support for the work reported in this manuscript without time limit. For all other items, the time frame for disclosure is the past 36 months.**

| | | **Name all entities with whom you have this relationship or indicate none (add rows as needed)** | **Specifications/Comments (e.g., if payments were made to you or to your institution)** |
|---|---|---|---|
| | | **Time frame: Since the initial planning of the work** | |
| 1 | All support for the present manuscript (e.g., funding, provision of study materials, medical writing, article processing charges, etc.) **No time limit for this item.** | ___X None | |
| | | **Time frame: past 36 months** | |
| 2 | Grants or contracts from any entity (if not indicated in item #1 above). | ____X None | |
| 3 | Royalties or licenses | ____ X None | |

| | | |
|---|---|---|
| 4 | Consulting fees | ____X None |
| 5 | Payment or honoraria for lectures, presentations, speakers bureaus, manuscript writing or educational events | ____ X None |
| 6 | Payment for expert testimony | ____X None |
| 7 | Support for attending meetings and/or travel | ____X None |
| 8 | Patents planned, issued or pending | ____ X None |
| 9 | Participation on a Data Safety Monitoring Board or Advisory Board | ____ X None |
| 10 | Leadership or fiduciary role in other board, society, committee or advocacy group, paid or unpaid | ____ X None |
| 11 | Stock or stock options | ____ X None |
| 12 | Receipt of equipment, materials, drugs, medical writing, gifts or other services | ____ X None |
| 13 | Other financial or non-financial interests | ____ X None |

**Please place an "X" next to the following statement to indicate your agreement:**

**_X__ I certify that I have answered every question and have not altered the wording of any of the questions on this form.**

**ICMJE DISCLOSURE FORM**

**Date:** 15 July 2021
**Your Name:** Valeria Ricotti
**Manuscript Title:** Investigating the Role of Dystrophin Isoform Deficiency in Motor Function in Duchenne Muscular Dystrophy
**Manuscript number (if known):**_______________________________________________________________

**In the interest of transparency, we ask you to disclose all relationships/activities/interests listed below that are related to the content of your manuscript. "Related" means any relation with for-profit or not-for-profit third parties whose interests may be affected by the content of the manuscript. Disclosure represents a commitment to transparency and does not necessarily indicate a bias. If you are in doubt about whether to list a relationship/activity/interest, it is preferable that you do so.**

**The following questions apply to the author's relationships/activities/interests as they relate to the current manuscript only.**

**The author's relationships/activities/interests should be defined broadly. For example, if your manuscript pertains to the epidemiology of hypertension, you should declare all relationships with manufacturers of antihypertensive medication, even if that medication is not mentioned in the manuscript.**

**In item #1 below, report all support for the work reported in this manuscript without time limit. For all other items, the time frame for disclosure is the past 36 months.**

| | | **Name all entities with whom you have this relationship or indicate none (add rows as needed)** | **Specifications/Comments (e.g., if payments were made to you or to your institution)** |
|---|---|---|---|
| | | **Time frame: Since the initial planning of the work** | |
| 1 | All support for the present manuscript (e.g., funding, provision of study materials, medical writing, article processing charges, etc.) **No time limit for this item.** | ___X None | Co-founder, EVP and CMO of DiNAQOR, shareholder of Solid Biosciences |
| | | **Time frame: past 36 months** | |
| 2 | Grants or contracts from any entity (if not indicated in item #1 above). | _____X None | |
| 3 | Royalties or licenses | _____ X None | |

| | | | |
|---|---|---|---|
| 4 | Consulting fees | ____X None | |
| 5 | Payment or honoraria for lectures, presentations, speakers bureaus, manuscript writing or educational events | ____ X None | |
| 6 | Payment for expert testimony | ____X None | |
| 7 | Support for attending meetings and/or travel | ____X None | |
| 8 | Patents planned, issued or pending | ____ X None | |
| 9 | Participation on a Data Safety Monitoring Board or Advisory Board | ____ X None | |
| 10 | Leadership or fiduciary role in other board, society, committee or advocacy group, paid or unpaid | ____ X None | |
| 11 | Stock or stock options | ____Yes | Shareholder of Solid Biosciences |
| 12 | Receipt of equipment, materials, drugs, medical writing, gifts or other services | ____ X None | |
| 13 | Other financial or non-financial interests | ____ Yes | Co-founder, EVP and CMO of DiNAQOR, |

**Please place an "X" next to the following statement to indicate your agreement:**

**X I certify that I have answered every question and have not altered the wording of any of the questions on this form.**

### ICMJE DISCLOSURE FORM

**Date:**___10th July 2021______________________________

**Your Name:**_____Yasumasa Hashimoto______________________________

**Manuscript Title:** Investigating the Role of Dystrophin Isoform Deficiency in Motor Function in Duchenne Muscular Dystrophy

**Manuscript number (if known):**______________________________

**In the interest of transparency, we ask you to disclose all relationships/activities/interests listed below that are related to the content of your manuscript. "Related" means any relation with for-profit or not-for-profit third parties whose interests may be affected by the content of the manuscript. Disclosure represents a commitment to transparency and does not necessarily indicate a bias. If you are in doubt about whether to list a relationship/activity/interest, it is preferable that you do so.**

**The following questions apply to the author's relationships/activities/interests as they relate to the current manuscript only.**

**The author's relationships/activities/interests should be defined broadly. For example, if your manuscript pertains to the epidemiology of hypertension, you should declare all relationships with manufacturers of antihypertensive medication, even if that medication is not mentioned in the manuscript.**

**In item #1 below, report all support for the work reported in this manuscript without time limit. For all other items, the time frame for disclosure is the past 36 months.**

| | | **Name all entities with whom you have this relationship or indicate none (add rows as needed)** | **Specifications/Comments (e.g., if payments were made to you or to your institution)** |
|---|---|---|---|
| | | **Time frame: Since the initial planning of the work** | |
| 1 | All support for the present manuscript (e.g., funding, provision of study materials, medical writing, article processing charges, etc.) **No time limit for this item.** | ____ None | |
| | | **Time frame: past 36 months** | |
| 2 | Grants or contracts from any entity (if not indicated in item #1 above). | ____ None | |
| 3 | Royalties or licenses | ____ None | |

| | | |
|---|---|---|
| 4 | Consulting fees | ____ None |
| 5 | Payment or honoraria for lectures, presentations, speakers bureaus, manuscript writing or educational events | ____ None |
| 6 | Payment for expert testimony | ____ None |
| 7 | Support for attending meetings and/or travel | ____ None |
| 8 | Patents planned, issued or pending | ____ None |
| 9 | Participation on a Data Safety Monitoring Board or Advisory Board | ____ None |
| 10 | Leadership or fiduciary role in other board, society, committee or advocacy group, paid or unpaid | ____ None |
| 11 | Stock or stock options | ____ None |
| 12 | Receipt of equipment, materials, drugs, medical writing, gifts or other services | ____ None |
| 13 | Other financial or non-financial interests | ____ None |

**Please place an "X" next to the following statement to indicate your agreement:**

**_X_ I certify that I have answered every question and have not altered the wording of any of the questions on this form.**

**ICMJE DISCLOSURE FORM**

**Date:12 07 2021**________________________________________________________________________________
**Your Name: Silvia Torelli**____________________________________________________________________________
**Manuscript Title:** Investigating the Role of Dystrophin Isoform Deficiency in Motor Function in Duchenne Muscular Dystrophy
**Manuscript number (if known):**_________________________________________________________________

**In the interest of transparency, we ask you to disclose all relationships/activities/interests listed below that are related to the content of your manuscript. "Related" means any relation with for-profit or not-for-profit third parties whose interests may be affected by the content of the manuscript. Disclosure represents a commitment to transparency and does not necessarily indicate a bias. If you are in doubt about whether to list a relationship/activity/interest, it is preferable that you do so.**

**The following questions apply to the author's relationships/activities/interests as they relate to the current manuscript only.**

**The author's relationships/activities/interests should be defined broadly. For example, if your manuscript pertains to the epidemiology of hypertension, you should declare all relationships with manufacturers of antihypertensive medication, even if that medication is not mentioned in the manuscript.**

**In item #1 below, report all support for the work reported in this manuscript without time limit. For all other items, the time frame for disclosure is the past 36 months.**

| | | **Name all entities with whom you have this relationship or indicate none (add rows as needed)** | **Specifications/Comments (e.g., if payments were made to you or to your institution)** |
|---|---|---|---|
| | | **Time frame: Since the initial planning of the work** | |
| 1 | All support for the present manuscript (e.g., funding, provision of study materials, medical writing, article processing charges, etc.) **No time limit for this item.** | ____ None | |
| | | **Time frame: past 36 months** | |
| 2 | Grants or contracts from any entity (if not indicated in item #1 above). | ____ None | |
| 3 | Royalties or licenses | ____ None | |

| | | |
|---|---|---|
| 4 | Consulting fees | _____ None |
| 5 | Payment or honoraria for lectures, presentations, speakers bureaus, manuscript writing or educational events | _____ None |
| 6 | Payment for expert testimony | _____ None |
| 7 | Support for attending meetings and/or travel | _____ None |
| 8 | Patents planned, issued or pending | _____ None |
| 9 | Participation on a Data Safety Monitoring Board or Advisory Board | _____ None |
| 10 | Leadership or fiduciary role in other board, society, committee or advocacy group, paid or unpaid | _____ None |
| 11 | Stock or stock options | _____ None |
| 12 | Receipt of equipment, materials, drugs, medical writing, gifts or other services | _____ None |
| 13 | Other financial or non-financial interests | _____ None |

**Please place an "X" next to the following statement to indicate your agreement:**

**__X_ I certify that I have answered every question and have not altered the wording of any of the questions on this form.**

### ICMJE DISCLOSURE FORM

**Date:___13th July 2021**
**Your Name:__ Kate Maresh**
**Manuscript Title:** Investigating the Role of Dystrophin Isoform Deficiency in Motor Function in Duchenne Muscular Dystrophy
**Manuscript number (if known):________________________________________________________________**

**In the interest of transparency, we ask you to disclose all relationships/activities/interests listed below that are related to the content of your manuscript. "Related" means any relation with for-profit or not-for-profit third parties whose interests may be affected by the content of the manuscript. Disclosure represents a commitment to transparency and does not necessarily indicate a bias. If you are in doubt about whether to list a relationship/activity/interest, it is preferable that you do so.**

**The following questions apply to the author's relationships/activities/interests as they relate to the current manuscript only.**

**The author's relationships/activities/interests should be defined broadly. For example, if your manuscript pertains to the epidemiology of hypertension, you should declare all relationships with manufacturers of antihypertensive medication, even if that medication is not mentioned in the manuscript.**

**In item #1 below, report all support for the work reported in this manuscript without time limit. For all other items, the time frame for disclosure is the past 36 months.**

| | | **Name all entities with whom you have this relationship or indicate none (add rows as needed)** | **Specifications/Comments (e.g., if payments were made to you or to your institution)** |
|---|---|---|---|
| **Time frame: Since the initial planning of the work** | | | |
| 1 | All support for the present manuscript (e.g., funding, provision of study materials, medical writing, article processing charges, etc.) **No time limit for this item.** | ____ None | |
| **Time frame: past 36 months** | | | |
| 2 | Grants or contracts from any entity (if not indicated in item #1 above). | Great Ormond Street Hospital Children's Charity; MRC Centre for Neuromuscular Diseases, Queen Square, London. | Payment to my institution. |

| | | |
|---|---|---|
| 3 | Royalties or licenses | ____ None |
| 4 | Consulting fees | ____ None |
| 5 | Payment or honoraria for lectures, presentations, speakers bureaus, manuscript writing or educational events | ____ None |
| 6 | Payment for expert testimony | ____ None |
| 7 | Support for attending meetings and/or travel | ____ None |
| 8 | Patents planned, issued or pending | ____ None |
| 9 | Participation on a Data Safety Monitoring Board or Advisory Board | ____ None |
| 10 | Leadership or fiduciary role in other board, society, committee or advocacy group, paid or unpaid | ____ None |
| 11 | Stock or stock options | ____ None |
| 12 | Receipt of equipment, materials, drugs, medical writing, gifts or other services | ____ None |
| 13 | Other financial or non-financial interests | ____ None |

**Please place an "X" next to the following statement to indicate your agreement:**

**__X_  I certify that I have answered every question and have not altered the wording of any of the questions on this form.**

**ICMJE DISCLOSURE FORM**

**Date:**___12/07/21_______________________________________________________________________
**Your Name:**____Deborah Ridout_____________________________________________________________
**Manuscript Title:** Investigating the Role of Dystrophin Isoform Deficiency in Motor Function in Duchenne Muscular Dystrophy
**Manuscript number (if known):**_____________________________________________________________________

**In the interest of transparency, we ask you to disclose all relationships/activities/interests listed below that are related to the content of your manuscript. "Related" means any relation with for-profit or not-for-profit third parties whose interests may be affected by the content of the manuscript. Disclosure represents a commitment to transparency and does not necessarily indicate a bias. If you are in doubt about whether to list a relationship/activity/interest, it is preferable that you do so.**

**The following questions apply to the author's relationships/activities/interests as they relate to the current manuscript only.**

**The author's relationships/activities/interests should be defined broadly. For example, if your manuscript pertains to the epidemiology of hypertension, you should declare all relationships with manufacturers of antihypertensive medication, even if that medication is not mentioned in the manuscript.**

**In item #1 below, report all support for the work reported in this manuscript without time limit. For all other items, the time frame for disclosure is the past 36 months.**

| | | **Name all entities with whom you have this relationship or indicate none (add rows as needed)** | **Specifications/Comments (e.g., if payments were made to you or to your institution)** |
|---|---|---|---|
| | | **Time frame: Since the initial planning of the work** | |
| 1 | All support for the present manuscript (e.g., funding, provision of study materials, medical writing, article processing charges, etc.) **No time limit for this item.** | ____ None | |
| | | **Time frame: past 36 months** | |
| 2 | Grants or contracts from any entity (if not indicated in item #1 above). | ____ None | |
| 3 | Royalties or licenses | ____ None | |

| | | |
|---|---|---|
| 4 | Consulting fees | ____ None |
| 5 | Payment or honoraria for lectures, presentations, speakers bureaus, manuscript writing or educational events | ____ None |
| 6 | Payment for expert testimony | ____ None |
| 7 | Support for attending meetings and/or travel | ____ None |
| 8 | Patents planned, issued or pending | ____ None |
| 9 | Participation on a Data Safety Monitoring Board or Advisory Board | ____ None |
| 10 | Leadership or fiduciary role in other board, society, committee or advocacy group, paid or unpaid | ____ None |
| 11 | Stock or stock options | ____ None |
| 12 | Receipt of equipment, materials, drugs, medical writing, gifts or other services | ____ None |
| 13 | Other financial or non-financial interests | ____ None |

**Please place an "X" next to the following statement to indicate your agreement:**

**_X_ I certify that I have answered every question and have not altered the wording of any of the questions on this form.**

Note to medRxiv – When the majority of the COI disclosures were completed, the manuscript title was ‘Investigating the Role of Dystrophin Isoform Deficiency in Motor Function in Duchenne Muscular Dystrophy’, however, prior to submission the title was changed to ‘Duchenne muscular dystrophy patients lacking the dystrophin isoforms Dp140 and Dp71 and mouse models lacking Dp140 have a more severe motor phenotype’
